# Regression calibration of self-reported mobile phone use to optimize quantitative risk estimation in the COSMOS study

**DOI:** 10.1101/2023.02.28.23286424

**Authors:** Marije Reedijk, Lützen Portengen, Anssi Auvinen, Katja Kojo, Sirpa Heinävaara, Maria Feychting, Giorgio Tettamanti, Lena Hillert, Paul Elliott, Mireille B Toledano, Rachel B Smith, Joël Heller, Joachim Schüz, Isabelle Deltour, Aslak Harbo Poulsen, Christoffer Johansen, Robert Verheij, Petra Peeters, Matti Rookus, Eugenio Traini, Anke Huss, Hans Kromhout, Roel Vermeulen, COSMOS Study Group

**Affiliations:** University of Utrecht, Institute for Risk Assessment Sciences, Utrecht, the Netherlands; Julius Center for Health Sciences and Primary Care, University Medical Center Utrecht (UMCU), Utrecht, the Netherlands; Tampere University, Faculty of Social Sciences, Tampere, Finland’; STUK – Radiation and Nuclear Safety Authority, Vantaa, Finland; Cancer Society of Finland/Finnish Cancer Registry, Helsinki, Finland; Karolinska Institutet, Institute of Environmental Medicine, Stockholm, Sweden; MRC Centre for Environment and Health, Imperial College London, School of Public Health, Department of Epidemiology and Biostatistics, London, UK; National Institute for Health and Care Research Health Protection Research Unit in Chemical and Radiation Threats and Hazards, Imperial College London, School of Public Health, Department of Epidemiology and Biostatistics, London, UK; Mohn Centre for Children’s Health and Wellbeing, School of Public Health, Imperial College London, London, UK; International Agency for Research on Cancer (IARC/WHO), Environment and Lifestyle Epidemiology Branch, Lyon, France; Danish Cancer Society Research Center, Copenhagen, Denmark; Danish Cancer Society Research Center, Copenhagen, Denmark; CASTLE Cancer Late Effect Research Oncology Clinic, Center for Surgery and Cancer, Rigshospitalet, Copenhagen, Denmark.; Netherlands Institute for Health Services Research (NIVEL), Utrecht, the Netherlands; The Netherlands Cancer Institute, Antoni van Leeuwenhoek Hospital, Amsterdam, The Netherlands

## Abstract

The Cohort Study of Mobile Phone Use and Health (COSMOS) study has repeatedly collected both self-reported and operator-recorded data on mobile phone use. Assessing health effects using self-reported information only is prone to measurement error, but operator data were available prospectively for only part of the study population and did not cover past mobile phone use. To optimize the available data and reduce bias, we evaluated different statistical approaches for constructing mobile phone exposure histories within COSMOS. We evaluated and compared the performance of complete case-analysis, different regression calibration methods, and multiple imputation in a simulation study with a binary health outcome. We used self-reported and operator-recorded mobile phone call data collected at baseline (2007-2012) from participants in Denmark, Finland, the Netherlands, Sweden, and the UK. Parameter estimates obtained using regression calibration methods were associated with less bias and lower mean squared error than those obtained with complete-case analysis or multiple-imputation. Our simulation study showed that regression calibration methods resulted in more accurate estimation of the relation between mobile phone use and health outcomes, by combining self-reported data with objective operator- recorded data available for a subset of the participants.

## Background

Exposure measurement error is a threat to the validity of environmental epidemiological research, and may be especially important for studies that rely on self-reported exposure information [1–3]. Systematic and random measurement error may result in substantial bias and loss of precision in estimated exposure-outcome relations [1, 3]. Validation studies have shown substantial error in self-reported estimates of mobile phone use [4–10]. Measurement error correction methods, such as regression calibration (RC), have been widely used in nutritional epidemiology [2,11–13], and are increasingly used in environmental epidemiology [3,14–18].

For mobile phone use, multiple bias modelling has been used in the Interphone Study [19], bias correction in the CEFALO study [6], and regression calibration by Tokola et al. [20]. Redmayne et al. developed a measurement error correction method for the number of weekly text messages [16]. The prospective *COhort Study of MObile phone uSe and health* (COSMOS) [9, 21] of more than 310,000 participants from 6 countries aimed to collect both self-reported and operator- recorded data on mobile phone use. Operator data could not be obtained for all participants, e.g. because subscriptions were held by employers, prepaid service, data sharing issues, or because participants did not consent to the provision of their mobile traffic data to the researchers [21].

Deciding which information to use for the primary analyses of exposure-outcome associations within COSMOS requires a careful trade-off between the available data sources on mobile phone use[10]. Use of operator-recorded information alone may lead both to imprecision in estimates, because data are available for only a subset of participants, as well as bias if data are not missing at random. Meanwhile use of more readily obtained (but error-prone) self-reported mobile phone data is likely to result in biased risk estimates and precision loss due to both systematic and random measurement errors.

Here we combined self-reported and operator-recorded mobile phone use data aiming to obtain acceptable precision of exposure-outcome estimates while minimizing the bias. We implemented regression calibration (RC) [3,22,23], complete-case (CC) analyses and multiple imputation (MI) [24] in a simulation study, comparing both bias and precision within and across methods, to optimize quantitative risk estimation in the COSMOS study.

## Methods

### Study design

We designed our simulation study using ‘real-world’ data from the COSMOS study, which has been described in detail elsewhere [21,25,26]. Briefly, information on mobile phone use was collected for participants aged 18 and over from 2007 onwards in six countries: Denmark, Finland, France, the Netherlands, Sweden, and the United Kingdom (UK) [21,25,26]. In Denmark, Finland, and Sweden, potential participants were selected using stratified random sampling from subscribers of major mobile phone network operators. In the Netherlands, recruitment was from the general population and a nurses cohort [26]. In the UK, approximately 65% of participants were recruited by stratified random sampling from subscribers of major mobile phone network operators, while 35% of participants were recruited from the electoral register [25] (Supplementary Table A1). Ethics committees reviewed and approved the study protocol in Denmark (Danish Data Protection Agency, J.nr: 2007-41-0856 and J.nr: 2005-41- 5533), Finland (Pirkanmaa Hospital District, tracking numbers R04179 and R09105), Sweden (Regional Ethical Review Board in Stockholm, 2007/1285–31/5, 2012/1608–32), and the UK (North West Haydock Research Ethics Committee, ref. 08/H1010/90). In the Netherlands, the Accredited Medical Research Ethics Committees (MRECs) assessed the protocol and found that the study did not require ethical review (protocol number 10/268C, 25/08/2010). Written or electronic informed consent to link to operator-recorded mobile phone use data was requested from each study participant.

### Questionnaire data

The COSMOS baseline questionnaire was administered between 2007-2012 either on paper or in most countries electronically. It was administered in 2019 in France, and the French data were therefore not considered in the current analysis. Key topics included mobile phone use, cordless phone use, use of other wireless devices, demographic and social characteristics and several self-reported health outcomes [21]. Characteristics considered as potential predictors of mobile phone use (or affecting self-report) were sex, age, marital status (living together, living apart, and not being in a relationship), educational level (elementary versus secondary school or higher), and employment status (active versus inactive).

### Self-reported mobile phone use

Self-reported duration of mobile phone use (REPORT) at baseline [21] was based on answers to the question: “*Over the last three months, on average, how much time per week did you spend talking on a mobile phone?”*. The following response options were provided: “< 5 min/week”, “5-29 min/week”, “30-59 min/week”, “1-3 hours/week”, “4-6 hours/week”, and “>6 hours/week”. In the Netherlands and the UK two further categories of call duration were included, “7-9 hours/week” and “10 or more hours/week”, but these were combined into “>6 hours/week” for the present analyses.

### Operator-recorded mobile phone use

Operator-recorded duration of mobile phone use (RECORD) was collected for all participants who provided consent and had a subscription under their own name. Operator-recorded data were used only when available for all mobile phones that were reported (up to two phones in Denmark, the Netherlands, Sweden, Finland, and up to three in the UK) and for at least three full months at the time the baseline questionnaire was administered. Participants with mobile phones that were also used by others were excluded. Operator-recorded data were available for 21% of participants in Denmark, 76% in Finland, 40% in the Netherlands, 63% in Sweden, and 76% in the UK.

Providers collected information on number and duration separately for outgoing and incoming calls. Across countries, outgoing calls made up 55-72% of the total number. Where the discrepancy between incoming and outgoing calls was large, this could suggest under-recording of incoming calls, possibly because calls between subscribers from the same provider were not always charged and may have gone unrecorded in some billing systems. For this reason, to evaluate the best RC method, we based our simulations on outgoing call duration only. Data for the three months at baseline were used to calculate average duration of calls in minutes per week.

### Simulation study

We compared results from different approaches that combine self-reported and operator-recorded mobile phone use: four variants of regression calibration (simple RC, Generalized Additive Model for Location, Shape, and Scale (GAMLSS)-based RC, direct RC, and inverse RC), a complete-case (CC) analysis and multiple imputation (MI) (Figure 1).

**Figure 1.**
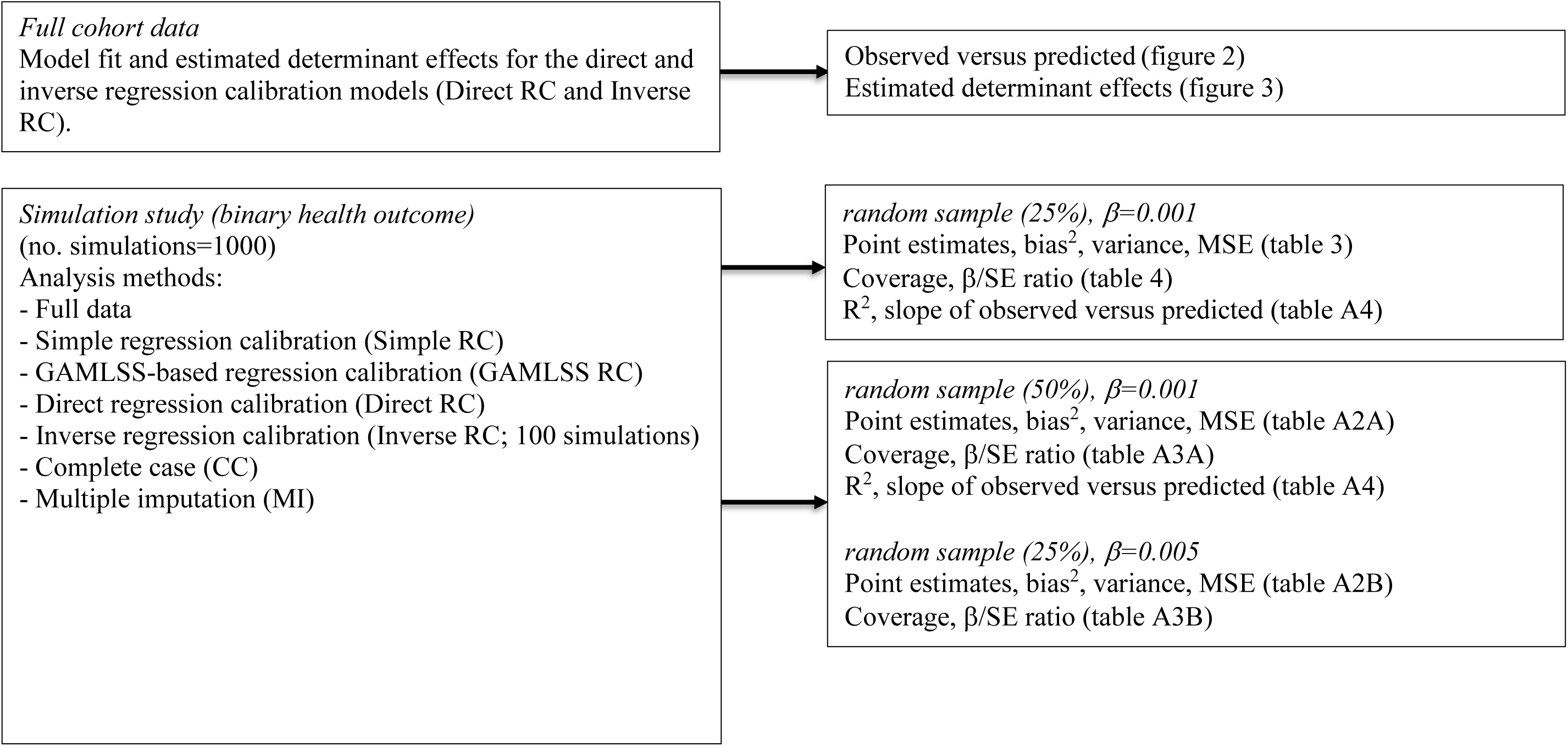
Roadmap to the main results from this paper. This includes evaluation of model fit and estimated determinant effects for RC models fitted to the available cohort data and results of the simulation study (including sensitivity analyses).

All simulations used data from the subset of participants for whom both self-reported and operator-recorded data were available. For each simulation, we assigned a binary health outcome with probability P(Yi=1) according to the following equation:

P(Yi=1) = exp(α + β*RECORDi)/(1+ exp(α + β*RECORDi))

for participant *i* with recorded call duration RECORDi (in minutes/week).

For our main analyses, the outcome was simulated using a slope coefficient (β) of 0.001 (i.e. assuming an Odds Ratio of exp(0.02)=1.02 per additional 20 minutes call-time per week) and with a balanced ratio of cases to non-cases (by tuning α). The value of 0.001 was chosen to provide a reasonably strong signal-to-noise ratio across the country-specific datasets [31].

Additional results using a slope coefficient (β) of 0.005 are presented in Supplementary Table A3B.

We randomly assigned 25% of the participants to the training set used to develop the regression calibration models and for fitting the complete-case model, while the remainder was used as a test set for fitting the “calibrated” health outcome model (mimicking the situation that provider data were missing for 75% of the population). Results obtained using 50% of participants as the training set are presented in Supplementary Table A4A.

The performance of each approach was evaluated by comparing the average estimated slope to the “true” slope coefficient used in the simulations; specifically, we estimated squared bias, variance, and mean squared error (MSE). We calculated 95% confidence intervals [95% CI] using bootstrapping to reflect Monte-Carlo error from using a limited number of simulations. We also estimated the coverage of 95% CIs calculated using the estimated standard error of the slope coefficient (Wald-type 95% CIs) to investigate whether these correctly accounted for the additional (sampling) variability. We calculated ratios of estimated slopes to estimated standard errors (β/SE) as a surrogate for statistical power and included results for the full data health model in the tables as an “ideal-case” reference method.

We used 1,000 simulations to evaluate performance for all approaches, except for the inverse RC method, where 100 simulations were used to avoid excessive running times. For the same reason, we used model-based (“naïve”) standard errors to obtain precision-weighted estimates, as we found these to be only marginally different from bootstrapped standard errors in a subset of 100 simulations.

### Full data and complete-case analysis

Logistic regression models were fitted to the simulated health outcome in the combined training and test datasets (“full data”) or the training set only (“complete-case”). Note that the “full data” option would not be available for COSMOS.

Operator-recorded mobile phone use was treated as gold standard for “*true”* mobile phone use, and the goal of our analyses was to estimate the slope coefficient (β) in the following health

g(E(Y)) = α + β.RECORD

where g is the logistic link function in the case of a binary outcome Y, and RECORD is operator- recorded mobile phone use.

### Regression calibration

Because RECORD is only available for a subset of the participants, we consider using self-reported mobile phone use (REPORT) as an (error-prone) proxy for operator-recorded use (RECORD) and consider different implementations of RC to correct for the measurement error [27]. All involve fitting the so-called “calibrated” health outcome model:

g(E(Y)) =α* + β*.E(RECORD|REPORT,Z)

where E(RECORD|REPORT,Z) is the expected value of “*true”* operator-recorded mobile phone use, conditional on REPORT and other covariates (Z).

We evaluated four different RC approaches, which differ by the amount of information used and in assumptions regarding the distribution of RECORD or on the relation between REPORT and RECORD. Our simple RC approach was the most basic, using the empirical average RECORD for each REPORT category. REPORT estimates appeared to follow an approximate log-normal distribution, and for the model-based approaches we indirectly estimated E(RECORD|REPORT,Z) using the relation between arithmetic mean (AM) and geometric mean (GM) plus geometric standard deviation (GSD) of a log-normally distributed variable (i.e. AM = GM*exp(log(GSD)^2^/2)). Our second approach, GAMLSS RC, used maximum likelihood in a generalized additive modeling framework to estimate both GM and GSD as a function of REPORT categories and covariates, allowing for non-linear effects of continuous covariates. Our third approach, direct RC, used regression splines to achieve the same flexibility in a Bayesian setting, but in addition relaxed distributional assumptions by modelling the residuals non-parametrically as a Dirichlet process mixture of (log)normal distributions, and with the AM estimated by averaging across draws from that distribution. Finally, our fourth approach, which we called inverse RC, was a Bayesian structural method and featured a probit regression model for REPORT categories using RECORD and all other covariates as predictor variables. By modelling the prior (conditional) distribution of RECORD as a Dirichlet process mixture of (log)normal distributions, we obtained estimates of RECORD conditional on REPORT and other covariates. The GAMLSS RC model was fitted using the R package gamlss [27], while the Bayesian RC models (direct, inverse) were fitted using the R package rjags [28].

Because health data in the COSMOS study will be available for most of the participants that contributed to the RECORD models, we followed the approach suggested by Spiegelman et al. [29] for studies that have an internal validation study. We combined the slope estimate from the complete-case model (i.e. the health model for the 25% of participants where RECORD was available) with that from the “calibrated” model fit to data from the remainder of the population (the test set), using inverse-precision weighting. Although robust standard errors for the slope estimate from the “calibrated” model could be obtained using bootstrapping, we did not do that here for computational reasons.

### Multiple imputation

Measurement error can also be regarded as a missing data problem [30], and we therefore considered multiple imputation (MI) in addition to the RC and complete-case analyses for the simulations. MI of operator-recorded phone use was performed by chained equations with fully conditional specification, using predictive mean matching as implemented in the R package mice [24]. Participant characteristics considered as predictors in the imputation model were the same as for the RC approaches, but also included the (simulated) health outcome (Y). We imputed a total of 10 different datasets and combined exposure slope coefficients from the health outcome model using Rubin’s rule [31].

### Model fit

We evaluated model fit for RC models using data from all participants with complete RECORD, REPORT, and the covariate data, for each country separately, by comparing predicted to observed average weekly call-time. We calculated the proportion of variance in observed call durations that could be explained by predicted call durations and evaluated the relation between predicted and observed call durations in detail by fitting a GAMLSS model that allowed for non-linear effects and non-homogeneous residual variance.

## Results

### Participant characteristics

Table 1 lists study attrition and personal characteristics for included participants. Complete information on covariates, REPORT (self-reported data), and RECORD (operator-recorded data) was available for <50% of participants for most countries, except for Finland and the UK. In the Netherlands, where sampling was from the general population and a nurses cohort, REPORT was not filled out by 33% of participants likely due to the high percentage of non-users. Slightly more women than men participated, except for the Netherlands, where most participants were women. There were notable differences in REPORT and RECORD between countries. RECORD was lowest in the Netherlands (GM [GSD] = 23.4 [2.6] minutes/week) and highest in Finland (81.5 [3.0] minutes/week).

**Table 1.**
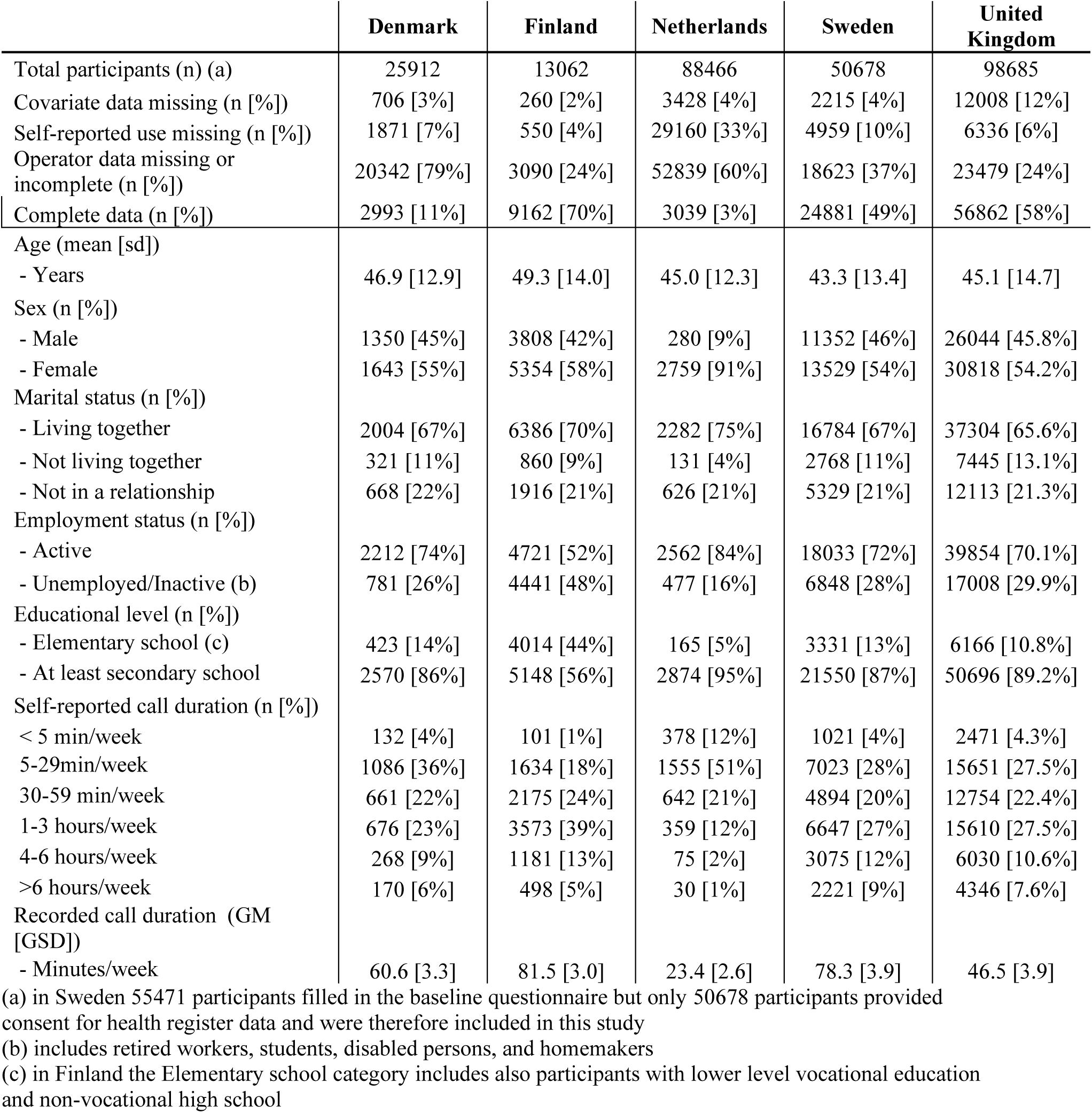
Overview of available data and personal characteristics for participants where both self-reported and operator-recorded data were available.

|  | Denmark | Finland | Netherlands | Sweden | United Kingdom |
| --- | --- | --- | --- | --- | --- |
| Total participants (n) (a) | 25912 | 13062 | 88466 | 50678 | 98685 |
| Covariate data missing (n [%]) | 706 [3%] | 260 [2%] | 3428 [4%] | 2215 [4%] | 12008 [12%] |
| Self-reported use missing (n [%]) | 1871 [7%] | 550 [4%] | 29160 [33%] | 4959 [10%] | 6336 [6%] |
| Operator data missing or incomplete (n [%]) | 20342 [79%] | 3090 [24%] | 52839 [60%] | 18623 [37%] | 23479 [24%] |
| Complete data (n [%]) | 2993 [11%] | 9162 [70%] | 3039 [3%] | 24881 [49%] | 56862 [58%] |
| Age (mean [sd]) |  |  |  |  |  |
| - Years | 46.9 [12.9] | 49.3 [14.0] | 45.0 [12.3] | 43.3 [13.4] | 45.1 [14.7] |
| Sex (n [%]) |  |  |  |  |  |
| - Male | 1350 [45%] | 3808 [42%] | 280 [9%] | 11352 [46%] | 26044 [45.8%] |
| - Female | 1643 [55%] | 5354 [58%] | 2759 [91%] | 13529 [54%] | 30818 [54.2%] |
| Marital status (n [%]) |  |  |  |  |  |
| - Living together | 2004 [67%] | 6386 [70%] | 2282 [75%] | 16784 [67%] | 37304 [65.6%] |
| - Not living together | 321 [11%] | 860 [9%] | 131 [4%] | 2768 [11%] | 7445 [13.1%] |
| - Not in a relationship | 668 [22%] | 1916 [21%] | 626 [21%] | 5329 [21%] | 12113 [21.3%] |
| Employment status (n [%]) |  |  |  |  |  |
| - Active | 2212 [74%] | 4721 [52%] | 2562 [84%] | 18033 [72%] | 39854 [70.1%] |
| - Unemployed/Inactive (b) | 781 [26%] | 4441 [48%] | 477 [16%] | 6848 [28%] | 17008 [29.9%] |
| Educational level (n [%]) |  |  |  |  |  |
| - Elementary school (c) | 423 [14%] | 4014 [44%] | 165 [5%] | 3331 [13%] | 6166 [10.8%] |
| - At least secondary school | 2570 [86%] | 5148 [56%] | 2874 [95%] | 21550 [87%] | 50696 [89.2%] |
| Self-reported call duration (n [%]) |  |  |  |  |  |
| < 5 min/week | 132 [4%] | 101 [1%] | 378 [12%] | 1021 [4%] | 2471 [4.3%] |
| 5-29min/week | 1086 [36%] | 1634 [18%] | 1555 [51%] | 7023 [28%] | 15651 [27.5%] |
| 30-59 min/week | 661 [22%] | 2175 [24%] | 642 [21%] | 4894 [20%] | 12754 [22.4%] |
| 1-3 hours/week | 676 [23%] | 3573 [39%] | 359 [12%] | 6647 [27%] | 15610 [27.5%] |
| 4-6 hours/week | 268 [9%] | 1181 [13%] | 75 [2%] | 3075 [12%] | 6030 [10.6%] |
| >6 hours/week | 170 [6%] | 498 [5%] | 30 [1%] | 2221 [9%] | 4346 [7.6%] |
| Recorded call duration (GM [GSD]) |  |  |  |  |  |
| - Minutes/week | 60.6 [3.3] | 81.5 [3.0] | 23.4 [2.6] | 78.3 [3.9] | 46.5 [3.9] |

(a) in Sweden 55471 participants filled in the baseline questionnaire but only 50678 participants provided consent for health register data and were therefore included in this study
(b) includes retired workers, students, disabled persons, and homemakers
(c) in Finland the Elementary school category includes also participants with lower level vocational education and non-vocational high school

The probability of having missing RECORD data depended on the value of REPORT data in all countries, but absolute differences were small and the direction across countries was inconsistent. A detailed comparison of participants with or without information on self-reported and operator- recorded mobile phone use and covariates per country is provided in Supplementary Table A2.

Figure 2 shows the distribution of RECORD for outgoing calls only, by country and categories of self-reported use. RECORD tended to be lower for participants from the Netherlands and the UK, even within categories of self-reported use, and this was most pronounced for higher REPORT categories.

**Figure 2.**
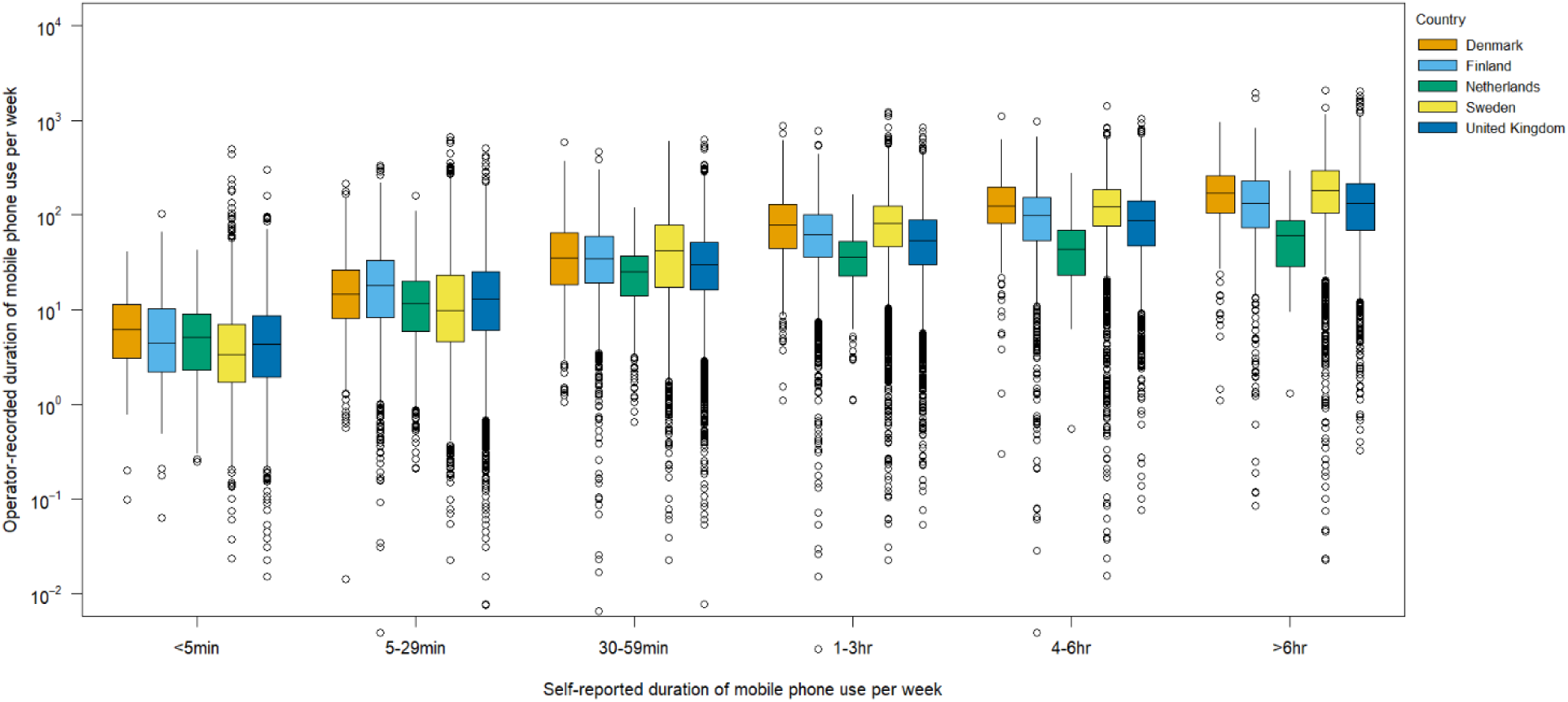
Outgoing operator-recorded mobile phone use by country and categories of self- reported use.

### Model fit and parameter estimates from RC models on the full data

Estimated covariate effects for the direct and inverse RC models fitted to the full cohort data, by country, are presented in the Supplementary material (Figures A1 and A2). Being in a relationship but not living together was a strong predictor for RECORD in all countries and was also associated with higher REPORT categories (conditional on RECORD) in the probit regression model for the inverse RC method (Figure A1). Older age was associated with lower RECORD and REPORT in most countries (Figure A2A/B). The underlying variable for REPORT in the probit regression model for the inverse RC method tended to decrease with age (Figure A2C), but increased with increasing RECORD (Figure A2D).

The relation between observed and predicted RECORD was close to linear for the direct RC model in most countries, but predictions in the high RECORD range tended to either underestimate (Finland, Sweden) or overestimate (Denmark, the Netherland, the UK) average observed values for the inverse RC model (Figure 3). Predictions from the GAMLSS-based RC model tended to be higher than observed in the high RECORD range for all countries (Figure A3). Differences in (in-sample) R^2^ between models were smaller than differences between countries, but were consistently larger for the direct than the inverse RC method. R^2^ was largest for RC models fitted to the Danish data (range 36.1-38.7%) and lowest for models fitted to the Finnish data (range 23.6-25.7%). Using any of the more complex RC methods resulted in slightly higher R^2 ‘^s (increase <2.6%) than the simple RC method (Supplementary Table A5). Country-specific regression calibrated estimates for the simple RC model based on outgoing call duration, and incoming and outgoing call duration combined, by category of self-report, are presented in Supplementary Table A6.

**Figure 3.**
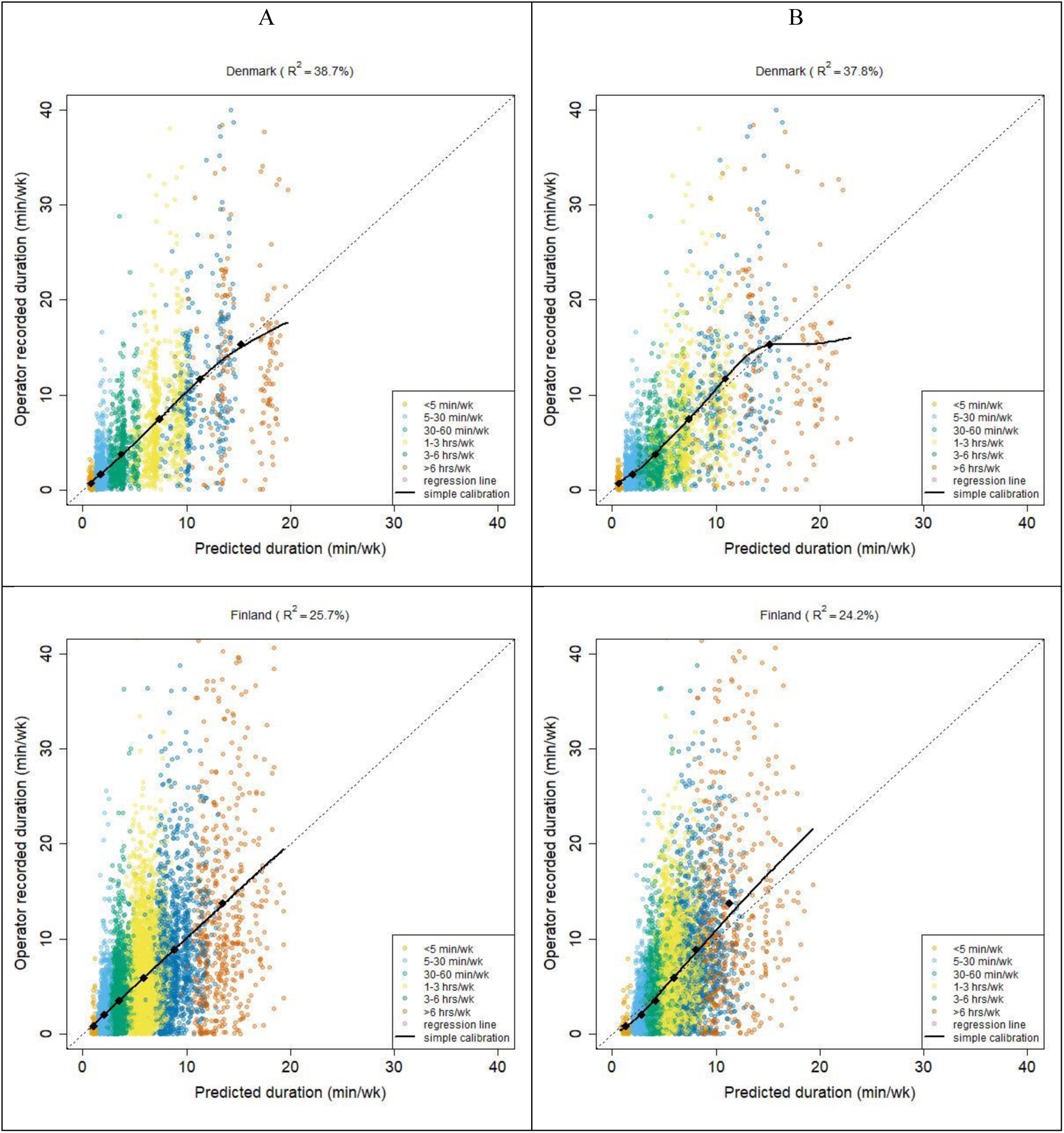
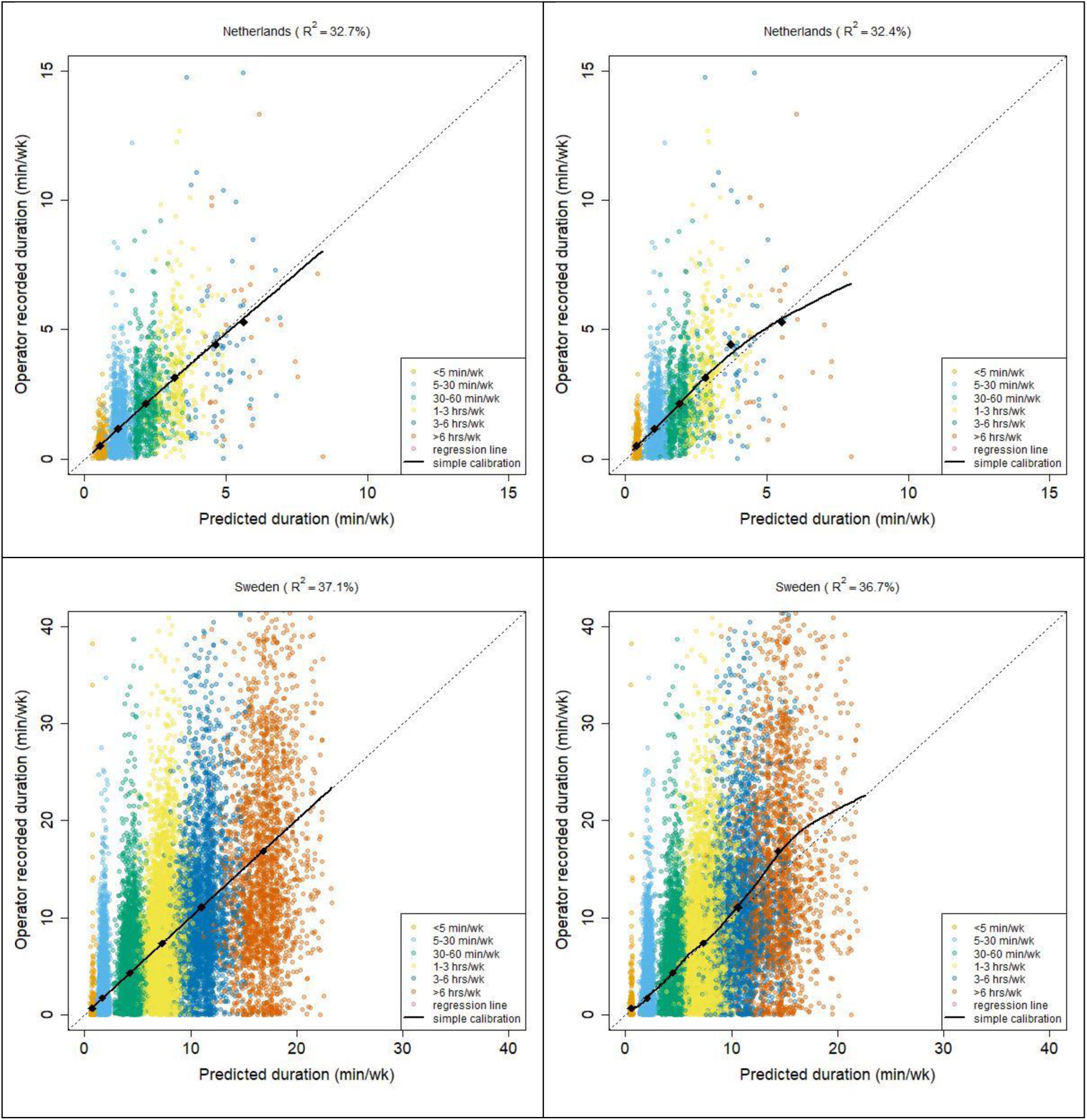
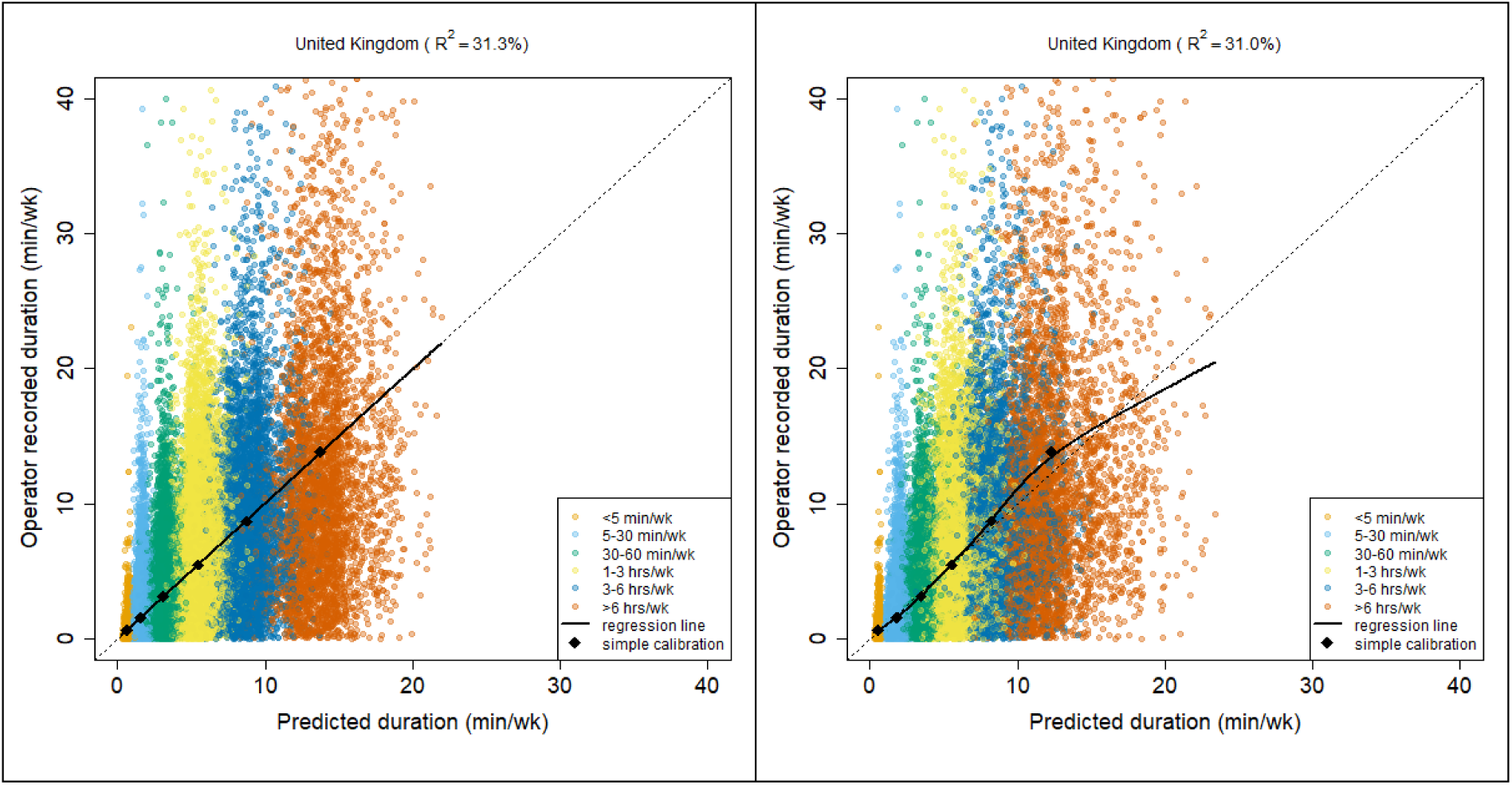
Observed versus predicted duration of outgoing calls by country for the direct (A) and inverse (B) RC approaches. The regression line was estimated allowing for a non-linear relation between observed and predicted values and allowing the (residual) variance in observed durations to depend on predicted duration using penalized splines (P-splines) as implemented in the gamlss software[29]. Note the different horizontal and vertical scales for the Netherlands.

### Performance of RC models on the simulated data

Simulation results are presented in Table 2. This table shows the (range of) estimated coefficients across simulations, as well as the squared bias and variance that contribute to total MSE. Using any of the RC approaches resulted in MSEs that were approximately 50% lower than those from CC analyses. MI was inferior to all RC approaches on average. Both simple RC and inverse RC performed relatively well on data from Finland, Sweden and the UK. MSEs were lower when a larger training set was used (50% of the sample; Supplementary Table A3A), especially for the inverse RC approach that performed well in almost all countries in both sets of simulations. Bias was a minor contributing factor to total MSE when the regression coefficient for RECORD used in the simulations was relatively small (0.001) (Supplementary Table A3B). Point estimates obtained using GAMLSS-based RC tended to suffer from relatively large downward bias.

**Table 2.**
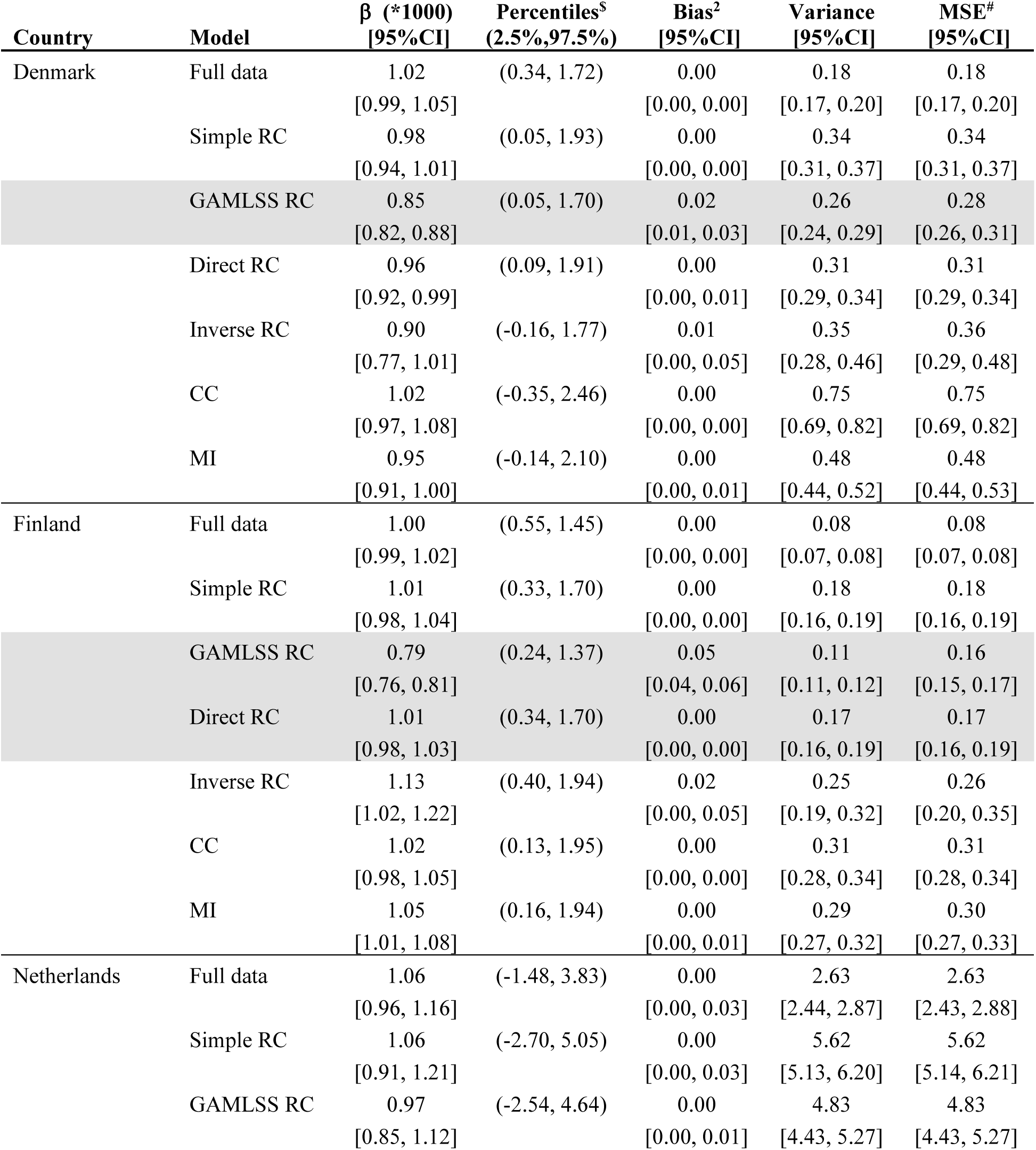
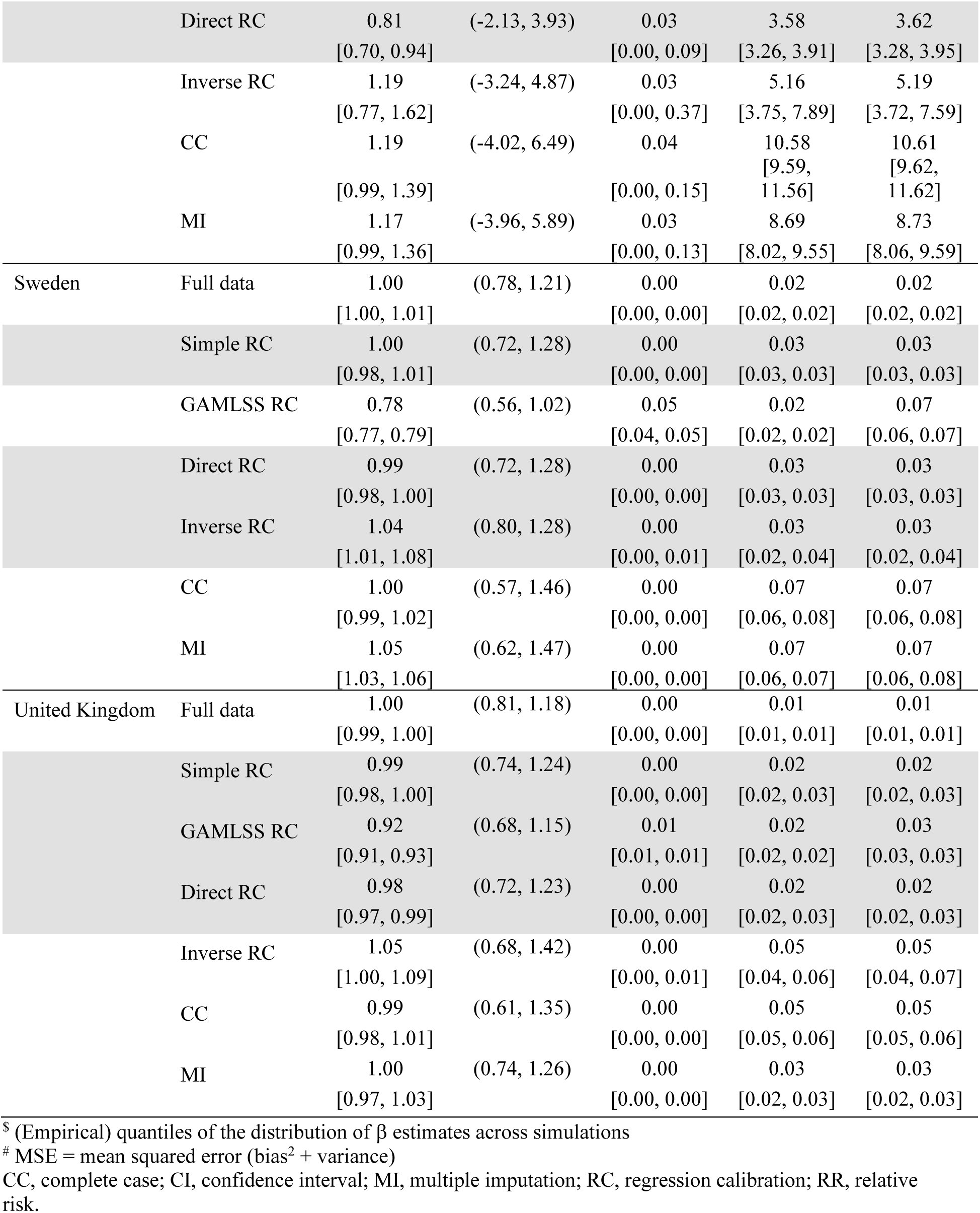
Point estimates, squared bias, variance and MSE for approaches based on the results from 1,000 simulations (100 simulations for the Inverse RC approach). Simulations used data from 25% of the participants with complete information on both self-reported and operator- recorded data as the training set and the remainder as test set. The outcome was simulated using a slope coefficient (β) of 0.001 (i.e. assuming an Odds Ratio of exp(0.1)=1.11 for each additional 100 minutes call-time per week) and with a balanced ratio of cases:non-cases. Bootstrapping was used to estimate 95%CIs for each statistic. The training set was used to fit the first stage (exposure) models for the RC approaches, the health model for the CC analysis, and the multiple imputation model. All second stage (health) models for the RC approaches were fitted to the test data only and results were precision-weighted with those from the CC approach before further analyses. The (non-full data) model that achieved lowest MSE and all models for which the estimated MSE fell within the 95%CI for that lowest MSE are highlighted in grey.

Coverage of Wald-type 95% confidence intervals based on estimated standard errors after inverse-precision weighting is shown in Table 3. Coverage was per expectation (95%) for the simple RC and direct RC methods, tended to be slightly lower for the inverse RC method (∼93%), but was much lower for the GAMLSS-based RC in some countries (e.g. 66% for Sweden). The average ratio of estimated slope (β) over standard error (SE) was highest for the direct RC or inverse RC methods for all countries, confirming higher statistical efficiency of these methods over alternative approaches.

**Table 3.**
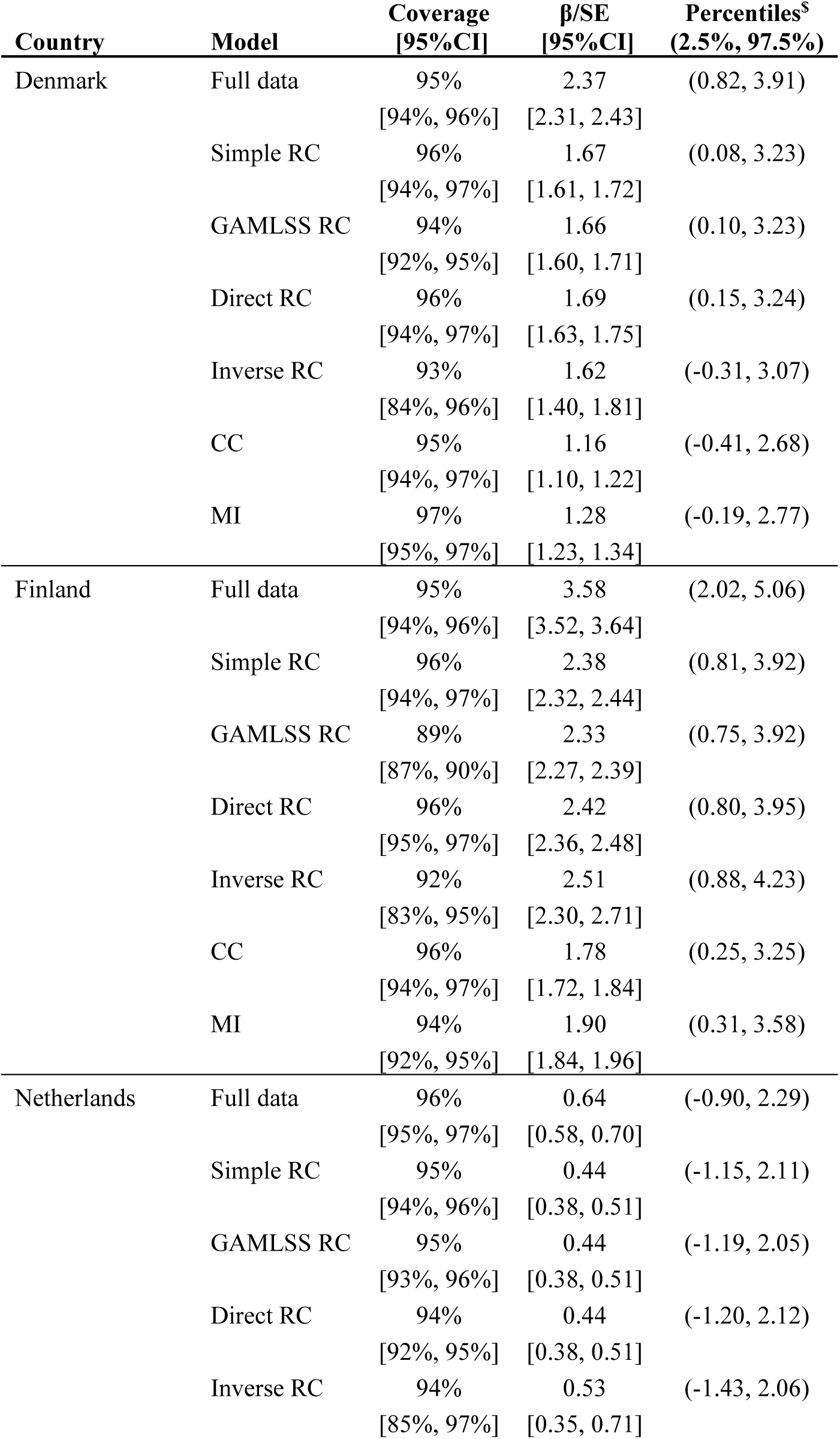
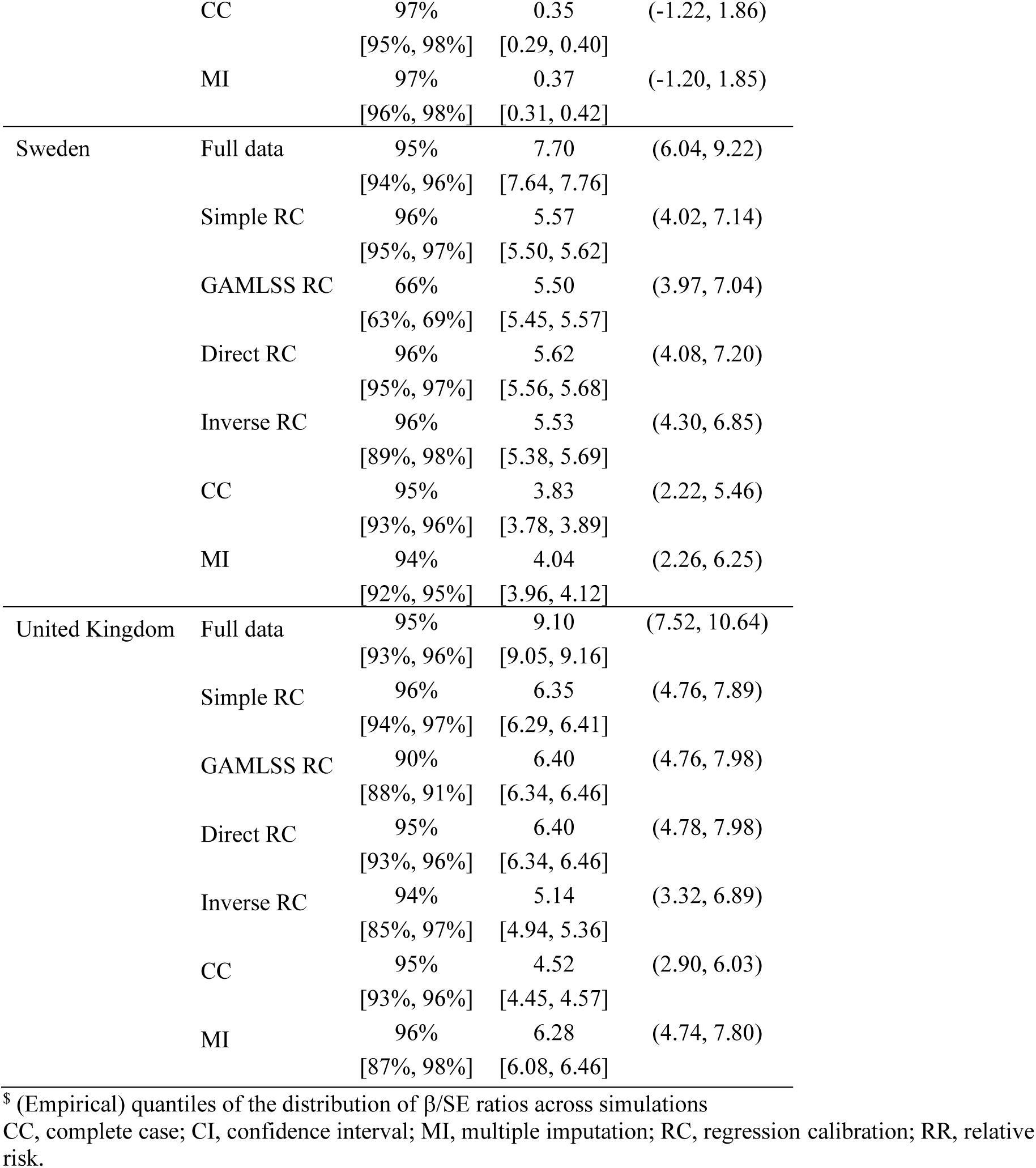
Coverage of 95%CI and ratio of slope estimate (β) over its standard error (SE) as a proxy for efficiency modelling approaches based on the results from 1,000 simulations (100 simulations for the Inverse RC approach). Simulations used data from 25% of the participants with complete information on both self-reported and operator-recorded data as the training set and the remainder as test set. The outcome was simulated using a slope coefficient (β) of 0.001 (i.e. assuming an Odds Ratio of exp(0.1)=1.11 for each additional 100 minutes call-time per week) and with a balanced ratio of cases:non-cases. Bootstrapping was used to estimate 95%CIs for each statistic.

## Discussion

We evaluated the performance of different approaches that combine operator-recorded mobile phone use and its more widely available, but error-prone proxy, self-reported use, in a simulation study using ‘real world’ COSMOS mobile phone use and covariate data with a simulated binary health outcome. We show that RC models, especially the simple and direct RC methods, provide a good balance between minimizing bias and maximizing precision and we therefore plan to use these methods in further analyses of the COSMOS data.

The results indicate that bias was a minor contributing factor to MSE of estimated slopes, when health outcomes were simulated under the assumption that the odds of disease increased by 2% for each additional 20 minutes of mobile phone call-time per week. Bias became more prominent when outcomes were simulated assuming a stronger exposure-outcome relation. The direction of bias (upwards or downwards) varied by country and RC approach. RC is expected to produce unbiased point estimates only for linear models [32], but our simulations indicate that bias was still limited for logistic regression models when the effect of (error-prone) reported use was not very strong or measurement error was relatively low by comparison [22,33,34], even with heteroscedastic error.

Total MSE of estimated slope coefficients were lowest for the full-data analyses and highest for the CC and MI methods, with relatively minor differences between different RC approaches. Among RC approaches, bias tended to be largest for the GAMLSS-based method. The simple RC approach performed only slightly worse than approaches relying on more complex models. This may be explained by the fact that covariates did not explain much of the variation in RECORD once REPORT was considered, as suggested by the modest increase in average R^2^ for more complex models when compared to simple RC. An advantage of using simple RC is that no additional covariates need to be available, while any gain from using more covariates would be (partly) offset by the need to consider these covariates as potential confounders in the health-outcome model [35, 36]. Also, requiring data on covariates to include in the model reduces the available sample size because of missing covariate data. We did not investigate this trade-off in this study, where health outcomes were simulated using only RECORD, but this could be of practical relevance when some of the model predictors are also known risk factors for the health outcome. Our results were based on simulations that assumed a linear relation between RECORD and the log-odds of our simulated health-outcome. This may be a reasonable assumption in many cases, but note that more complicated calibration models are required if the goal is to use the calibrated exposure estimate in a non-parametric or polynomial regression model [37].

RC models were fitted using two-stage regression, and standard errors from the second stage model do not fully account for the uncertainty in exposure estimates from the first stage models. Standard errors could be estimated through (asymptotic) approximation or bootstrapping [22, 32], but for practical reasons we used model-based standard errors for the simulations in our analyses. Coverage of estimated 95% CIs was only slightly lower than expectation for most approaches, except for the GAMLSS-based RC, where bias in point estimates was substantial.

Simulated health outcomes were included in the MI models, but we did not consider fitting a full measurement error model that incorporates the health outcome regression [38]. Feedback between the health outcome model and the exposure and measurement error models could result in a more efficient analysis of the exposure-outcome relation, but could also result in bias amplification from model misspecification [39]. A full measurement error model would also require re-fitting the model separately for each health outcome or including all health outcomes in a single model, which is unattractive for a multi-center cohort study.

Other authors have suggested methods to either account for measurement error in self-reported mobile phone data or to assess the potential impact on study findings, but these too have their limitations. Redmayne et al.[16] used a Bayesian forecasting method to correct for measurement error in the self-reported number of text messages. Their method shows similarities to the inverse RC approach applied here, but their model did not include any covariates and self-reported use was on a continuous scale. Vergnaud et al. [40] imputed missing information on Terrestrial Truncated Radio (TETRA) use among police forces using machine learning techniques. They did not include health outcomes in their imputation model and primarily reported results based on personal exposure estimates averaged across 10,000 imputations, which made it similar to our GAMLSS-based RC method. They found, as we did, that this model tended to under-estimate use for higher exposures.

Complete information on covariates, self-reported phone use, and operator-recorded data was available for a third of participants (N=96,937, 35.0%), and was collected prior to disease occurrence. This addresses a major limitation of earlier case-control studies which relied on retrospective information on mobile phone use, although missing information could be associated with some of the health outcomes of interest. Operator data can also be problematic as they are collected by providers for billing or security reasons rather than scientific purposes. In some countries, calls made between a participant and another caller where both were using the same provider were registered as outgoing calls only, so some incoming calls may be missing. We therefore based our simulation study on outgoing call duration, even though we may choose to use calibrated call duration estimates based on combined incoming and outgoing call duration data in COSMOS.

Collection of operator-recorded data within COSMOS will likely become more difficult over time due to attrition and participants changing network operators. Self-reported information on mobile phone use will therefore remain important [10], but may require the use of structural measurement error models to account for time-trends during follow-up, ever-changing patterns in mobile phone use, and newer network technologies. Notably, there are limitations inherent in capturing evolving mobile phone technology changes over time that may impact the estimation of exposure-response relations, which cannot be addressed with our approach.

A major issue is how well mobile phone use predicts the exposure of interest, namely radiofrequency electromagnetic fields. While past validation studies have been carried out for the 2^nd^ mobile phone generation, showing fair agreement between amount of use and cumulated emission from the handset [4], fewer data are available on the predictive power of mobile phones of the 3^rd^, 4^th^ and 5^th^ generations that have and are being used by COSMOS participants.

## Conclusion

This study addressed an important concern in mobile phone research and more generally in environmental epidemiology: how to leverage self-reported exposure estimates that are often available but error-prone, with more objective measurements that may be obtained in only a subset of participants. Our simulation study indicated RC approaches may improve estimation of exposure-outcome relations between mobile phone use and health outcomes within COSMOS.

The prospective design and improved exposure assessment within COSMOS compared to that in previous case-control studies are expected to lead to more robust conclusions about possible health effects from use of mobile phones.

## Data Availability

All data produced in the present study are available upon reasonable request to the authors.

## Acknowledgements

We want to thank all the participants who have joined the COSMOS cohort study (http://www.thecosmosproject.org/). We thank mobile phone network operators in Denmark, Finland, France, the Netherlands, Sweden and the UK for allowing invitation of their subscribers and/or provision of operator traffic data.

COSMOS Study Group members not in the author list: Terhi Lampio, Susanna Lankinen, Turkka Näppilä and Taru Vehmasto, Tampere University, Faculty of Social Sciences, Tampere, Finland; Inka Pieterson, Institute for Risk Assessment Sciences (IRAS), Utrecht University, Utrecht, the Netherlands; David Muller, Margaret Douglass, and James Brook, Department of Epidemiology and Biostatistics, School of Public Health, Imperial College London; Mats Talbäck, Unit of Epidemiology, Institute of Environmental Medicine, Karolinska Institutet, Stockholm Sweden. We thank Anders Ahlbom, Institute of Environmental Medicine, Karolinska Institutet, Stockholm Sweden, for his essential role in commencing the COSMOS cohort collaboration.

PE is Director of the MRC Centre for Environment and Health supported by the Medical Research Council (MR/S019669/1). PE acknowledges funding from the NIHR Imperial Biomedical Research Centre, the NIHR Health Protection Research Unit in Chemical and Radiation Threats and Hazards (NIHR-200922) and the UK Dementia Research Institute supported by UK DRI Ltd which is funded by the UK Medical Research Council, Alzheimer’s Society and Alzheimer’s Research UK (UKDRI-5001). PE is associate director of Health Data Research UK-London which receives funding from a consortium led by the UK Medical Research Council. MBT’s Chair and RBS’s fellowship are supported by a donation from Marit Mohn to Imperial College London to support Population Child Health through the Mohn Centre for Children’s Health and Wellbeing. MR, LP, RV and HK were supported by The Netherlands Organization for Health Research (ZonMW) within the programme Electromagnetic Fields and Health Research, under grant numbers 85200001, 85500003 and 85800001. Where authors are identified as personnel of the International Agency for Research on Cancer/World Health Organization, the authors alone are responsible for the views expressed in this article and they do not necessarily represent the decisions, policy or views of the International Agency for Research on Cancer /World Health Organization.

