## Supplementary material for "Regression calibration of self-reported mobile phone use to optimize quantitative risk estimation in the COSMOS study": medRxiv_COSMOS regression calibration.supplementary.v14.docx

**Supplementary methods**
We start with describing the health outcome model used for the *complete case* (CC) and second stage analyses below, followed by a description of the different *regression calibration* (RC) approaches used to build the first stage models and the *multiple imputation* (MI) procedure.

*Health outcome model*
We regard the operator-recorded mobile phone use data as the gold standard for *“true”* mobile phone use, and the goal of our analyses is therefore to estimate the slope coefficient (β) in the following health outcome model:

g(E(Y)) = α + β.RECORD + γ.C	(eq. I)

where g is the logistic link function in case of a binary outcome Y, RECORD is operator-recorded mobile phone use, and C is a matrix of known or suspected confounders. Non-linear effects of either exposure or confounders can be accommodated using basis-expansions methods (f.i. using a cubic spline basis for RECORD or age). We did not include any scenarios that involve confounding in our simulations, so C is not included in the health outcome model we use, but it should be noted that predictors used in the first stage (exposure) models should be considered potential confounders by design[1].

*Complete case analysis*
For the *complete case* analysis, we fitted the health outcome model (eq. I) using data from participants in the training dataset only. Unless the number of participants without operator-recorded phone use information is low, this option is likely to be inefficient, resulting in high standard errors for the regression coefficient (β). When operator-recorded information is not missing completely at random (MCAR), complete case analysis could also result in biased estimation of this regression coefficient. For the simulations in this paper, random sampling was used to assign participants to either training or test datasets, and therefore the MCAR assumption holds and no bias is expected.

*Regression calibration approaches*
We consider self-reported mobile phone use (REPORT) as an (error-prone) proxy for operator-recorded use (RECORD) and consider four different implementations of *regression calibration*[2] (RC) to correct for measurement error. This involves fitting the so-called *“calibrated”* health outcome model:

g(E(Y)) = α^*^ + β^*^.E(RECORD|REPORT,Z)	+ γ^*^.C	(eq. II)

where E(RECORD|REPORT,Z) is the expected value of *“true”* operator-recorded mobile phone use, conditional on REPORT and other covariates (Z). This model may be used to adjust slope estimates from a health outcome model fitted to the error-prone exposure proxy, but adjusted slope estimates can also be obtained through two-stage regression, as we do here. In that case, the *“calibrated”* health outcome model (eq. II) is the second-stage model, while E(RECORD|REPORT,Z) is estimated from the regression of RECORD on REPORT and other covariates in the first-stage model using the training sample:

E(RECORD|REPORT,Z) = φ + η.REPORT + λ.Z	(eq. III)

Our first approach, simple regression calibration (simple RC), does not use any covariates and estimates E(RECORD|REPORT) by simply averaging operator-recorded mobile phone use (RECORD) across different levels of the REPORT categories.

For the other variants we chose Z to include sex, age, educational achievement, employment status, and marital status of the study participants. As for the health outcome model, non-linear effects of continuous covariates can be accommodated using basis expansion methods, and we use a three degrees of freedom cubic spline basis for age here as well. The distribution of RECORD is strongly skewed to the right, even withing REPORT categories, and we therefore chose to estimate the (conditional) mean of RECORD using models for log-transformed RECORD as follows:

E(RECORD|REPORT,Z) = exp(E(log(RECORD)|REPORT,Z) + Var(log(RECORD)|REPORT,Z)/2) (eq. IV)

with

E(log(RECORD)|REPORT,Z) = φ_log_ + η_log_.REPORT + λ_log_.Z	(eq. V)

Var(log(RECORD)|REPORT,Z) = exp(φ_var_ + η_var_.REPORT + λ_var_.Z)	(eq. VI)

Our second approach, which we call GAMLSS regression calibration (GAMLSS RC), uses software to fit generalized additive models for location, shape, and scale (GAMLSS) to estimate equations V and VI, and then uses equation IV to estimate E(RECORD|REPORT,Z).

A challenge in estimating equation IV using GAMLSS models is that a good estimate of Var(log(RECORD)|REPORT,Z) is critically important but that the (conditional) distribution of log(RECORD) is still noticably skewed across REPORT categories and covariates (Z). As an alternative we therefore used a non-parametric Bayesian approach to approximate the residual distribution by fitting a (truncated) Dirichlet process mixture of (log)normal distributions[3] to the residuals within each REPORT category and averaging across draws from that distribution. We used uninformative priors for the regression parameters and truncated the Dirichlet process to use no more than 5 mixture components per REPORT category to avoid excessive running times for the simulations, but allowed for more components (n=10) when fitting the model to actual cohort data. The models were fitted using the Bayesian modelling software JAGS[4].We call this (third) approach “direct” regression calibration (direct RC) to distinguish it from the next.

The GAMLSS-based RC and direct RC approaches outlined above ignore the causal relation between RECORD and REPORT, treating REPORT as a categorical predictor of RECORD instead (without introducing any constraints to guarantee monotonicity), so we considered ways to improve the model by imposing additional structure on the regression estimates, i.e. by explicitly modelling how RECORD and other covariates could influence self-reported phone use. This fourth approach, which we call inverse regression calibration (inverse RC) includes a probit regression model for the (ordinal) REPORT variable with an underlying (latent) variable (U*) that is affected by both RECORD and other covariates. The observed REPORT categories result from thresholding this variable, as follows:

E(U*) = υ.RECORD + ω.Z	(eq. VII)

with (τ_i_ < U* ≤ τ_i+1_) implying that REPORT = i

We allow for possible non-linear effects of RECORD and components of Z by using basis expansion methods as in the other models. To allow for missing (unobserved) RECORD variables, we estimate the distribution of RECORD semi-parametrically, as a truncated Dirichlet process mixture of normal distributions which we truncate at 5 components for the simulations (and 10 for fits to the full cohort data). To obtain estimates of RECORD conditional on categories of REPORT and other covariates from this model, we need to inverse the probit model, which can be done relatively easily within a Bayesian framework. We used flat priors for the regression parameters and fitted the models using JAGS[4].

Because health data are available for all participants that contribute to the first-stage model (i.e. have operator-recorded data), we follow the approach suggested by Spiegelman et al.[2] for studies that have an internal validation study. This approach consists of combining the slope estimate from the complete-case model fit to the internal validation data (the training set) with that from the second stage model (the “calibrated” health model) fit to the remainder of the data (the test set), using inverse-precision weighting. For the actual COSMOS analsysis, where it may be important to check for differences in exposure-response between subjects with or without operator-recorded data, one could assign the calibrated phone use data only to participants in the test set and use the operator-recorded phone use for participants in the training set to allow more direct testing of slope equality and other transportability checks.

*Multiple imputation*
Measurement error can be regarded as a missing data issue[5], and we therefore considered multiple imputation (MI) in addition to the complete-case and RC analyses for the simulations. MI of operator-recorded phone use was performed by chained equations (fully conditional specification) using predictive mean matching as implemented in the R package mice[6]. Participant characteristics considered as potential predictors in the imputation model were the same as for the Indirect RC approach (i.e. self-reported mobile phone use, sex, age, educational achievement, employment status, and marital status), but additionally included the (simulated) health outcome (Y). We allowed for a potential non-linear relation between age and RECORD by using a cubic spline basis expansion for age with 3 degrees of freedom. We imputed a total of 10 different datasets and combined exposure slope coefficients from the health outcome model using Rubin’s rule[7].

**Table A1.** Study design: participating countries, enrolment year, and recruitment strategy.

| **Country** | **Enrolment** | | **Recruitment strategy** | **Questionnaire** |
| --- | --- | --- | --- | --- |
|  | **Year** | **N** |  |  |
| **Denmark** | 2007-2009 | 25912 | Stratified random sampling from operator subscription data (a) | Paper |
| **Sweden** | 2008-2009 | 50678 | Stratified random sampling from operator subscription data (a) | Paper |
| **Finland** | 2009-2010 | 13062 | Stratified random sampling from operator subscription data (b) | Paper/Electronic |
| **UK** | 2009-2012 | 98685 | Stratified random sampling from operator subsctiption data (b) / General population sampling | Electronic |
| **Netherlands** | 2011-2012 | 88466 | General populaton sampling and  occupational population sampling (nurses) | Paper/ Electronic |

(a) Stratified random sampling from operator subscription data based on call time and age

(b) Stratified random sampling from operator subscription data based on call time, age, and sex

**Table A2**. Comparison of subject characteristics for participants in the COSMOS study that either had or did not have information available on subject covariate data, self-reported mobile phone use, and operator-recorded data on mobile phone use (duration of calls), by country. P values in the last column indicate statistical significance of differences between subjects with covariate and self-reported mobile phone use data that either had or did not have operator-recorded data available (the two penultimate columns) and is based on the chi-square test for categorical variables and the Wilcoxon rank-test for age and recorded call duration. Note that the category of subjects that were unemployed/inactive includes retired workers, students, disabled persons, and homemakers.

|  | **Denmark** | | | | |  |
| --- | --- | --- | --- | --- | --- | --- |
|  | Covariate data missing | Self-reported & operator-recorded data missing | Non-users or self-reported data missing | Operator-recorded data missing | Complete data available | P |
| Number of participants (n) | 706 | 1859 | 12 | 20342 | 2993 |  |
| Age |  |  |  |  |  |  |
| - No. observations | - | - | - | - | - |  |
| - Years (mean [sd]) | 50.9 [10.7] | 57.3 [9.7] | 53.4 [14.2] | 50.7 [11.2] | 46.9 [12.9] | <0.001 |
| Sex (n [%]) |  |  |  |  |  |  |
| - No. observations | - | - | - | - | - |  |
| - Male | 285 [40%] | 797 [43%] | 6 [50%] | 10504 [52%] | 1350 [45%] | <0.001 |
| - Female | 421 [60%] | 1062 [57%] | 6 [50%] | 9838 [48%] | 1643 [55%] |  |
| Marital status (n [%]) |  |  |  |  |  |  |
| - No. observations | 380 [54%] | - | - | - | - |  |
| - Living together | 215 [57%] | 1468 [79%] | 5 [42%] | 15712 [77%] | 2004 [67%] | <0.001 |
| - Not living together | 47 [12%] | 62 [3%] | 0 [0%] | 1308 [6%] | 321 [11%] |  |
| - Not in a relationship | 118 [31%] | 329 [18%] | 7 [58%] | 3322 [16%] | 668 [22%] |  |
| Employment status (n [%]) |  |  |  |  |  |  |
| - No. observations | 240 [34%] | - | - | - | - |  |
| - Active | 165 [69%] | 1062 [57%] | 8 [67%] | 15538 [76%] | 2212 [74%] | <0.001 |
| - Unemployed/Inactive | 75 [31%] | 797 [43%] | 4 [33%] | 4804 [24%] | 781 [26%] |  |
| Educational level (n [%]) |  |  |  |  |  |  |
| - No. observations | 586 [83%] | - | - | - | - |  |
| - Elementary school | 117 [20%] | 235 [13%] | 4 [33%] | 2608 [13%] | 423 [14%] | 0.05 |
| - At least secondary school | 469 [80%] | 1624 [87%] | 8 [67%] | 17734 [87%] | 2570 [86%] |  |
| Self-reported call duration (n [%]) |  |  |  |  |  |  |
| - No. observations | 647 [92%] | - | - | - | - |  |
| < 5 min/week | 49 [8%] | - | - | 1484 [7%] | 132 [4%] | <0.001 |
| 5-29min/week | 230 [36%] | - | - | 8057 [40%] | 1086 [36%] |  |
| 30-59 min/week | 131 [20%] | - | - | 4334 [21%] | 661 [22%] |  |
| 1-3 hours/week | 156 [24%] | - | - | 4184 [21%] | 676 [23%] |  |
| 4-6 hours/week | 50 [8%] | - | - | 1420 [7%] | 268 [9%] |  |
| >6 hours/week | 31 [5%] | - | - | 863 [4%] | 170 [6%] |  |
| Recorded call duration |  |  |  |  |  |  |
| - No. observations | 88 [12%] | - | - | - | - |  |
| - Minutes/week (GM [GSD]) | 73.4 [3.7] | - | 28.0 [4.1] | - | 60.6 [3.3] |  |

|  | **Finland** | | | | |  |
| --- | --- | --- | --- | --- | --- | --- |
|  | Covariate data missing | Self-reported & operator-recorded data missing | Non-users or self-reported data missing | Operator-recorded data missing | Complete data available | P |
| Number of participants (n) | 260 | 101 | 449 | 3090 | 9162 |  |
| Age |  |  |  |  |  |  |
| - No. observations | 258 [99%] | - | - | - | - |  |
| - Years (mean [sd]) | 55.3 [12.9] | 53.0 [13.6] | 53.6 [13.2] | 48.2 [13.1] | 49.3 [14.0] | <0.001 |
| Sex (n [%]) |  |  |  |  |  |  |
| - No. observations | - | - | - | - | - |  |
| - Male | 110 [42%] | 40 [40%] | 196 [44%] | 1529 [49%] | 3808 [42%] | <0.001 |
| - Female | 150 [58%] | 61 [60%] | 253 [56%] | 1561 [51%] | 5354 [58%] |  |
| Marital status (n [%]) |  |  |  |  |  |  |
| - No. observations | 208 [80%] | - | - | - | - |  |
| - Living together | 147 [71%] | 59 [58%] | 279 [62%] | 2253 [73%] | 6386 [70%] | <0.001 |
| - Not living together | 13 [6%] | 6 [6%] | 31 [7%] | 268 [9%] | 860 [9%] |  |
| - Not in a relationship | 48 [23%] | 36 [36%] | 139 [31%] | 569 [18%] | 1916 [21%] |  |
| Employment status (n [%]) |  |  |  |  |  |  |
| - No. observations | 123 [47%] | - | - | - | - |  |
| - Active | 46 [37%] | 48 [48%] | 187 [42%] | 2056 [67%] | 4721 [52%] | <0.001 |
| - Unemployed/Inactive | 77 [63%] | 53 [52%] | 262 [58%] | 1034 [33%] | 4441 [48%] |  |
| Educational level (n [%]) |  |  |  |  |  |  |
| - No. observations | 139 [53%] | - | - | - | - |  |
| - Elementary school | 80 [58%] | 45 [45%] | 231 [51%] | 1145 [37%] | 4014 [44%] | <0.001 |
| - At least secondary school | 59 [42%] | 56 [55%] | 218 [49%] | 1945 [63%] | 5148 [56%] |  |
| Self-reported call duration (n [%]) |  |  |  |  |  |  |
| - No. observations | - | - | - | - | - |  |
| < 5 min/week | 8 [3%] | - | - | 19 [1%] | 101 [1%] | <0.001 |
| 5-29min/week | 42 [16%] | - | - | 466 [15%] | 1634 [18%] |  |
| 30-59 min/week | 52 [20%] | - | - | 645 [21%] | 2175 [24%] |  |
| 1-3 hours/week | 83 [32%] | - | - | 1173 [38%] | 3573 [39%] |  |
| 4-6 hours/week | 28 [11%] | - | - | 482 [16%] | 1181 [13%] |  |
| >6 hours/week | 47 [18%] | - | - | 305 [10%] | 498 [5%] |  |
| Recorded call duration |  |  |  |  |  |  |
| - No. observations | 206 [79%] | - | - | - | - |  |
| - Minutes/week (GM [GSD]) | 84.2 [3.0] | - | 46.5 [4.6] | - | 81.5 [3.0] |  |

|  | **Netherlands** | | | | |  |
| --- | --- | --- | --- | --- | --- | --- |
|  | Covariate data missing | Self-reported & operator-recorded data missing | Non-users or self-reported data missing | Operator-recorded data missing | Complete data available | P |
| Number of participants (n) | 3428 | 28545 | 615 | 52839 | 3039 |  |
| Age |  |  |  |  |  |  |
| - No. observations | - | - | - | - | - |  |
| - Years (mean [sd]) | 64.6 [12.6] | 54.4 [11.8] | 49.4 [10.1] | 47.2 [11.9] | 45.0 [12.3] | <0.001 |
| Sex (n [%]) |  |  |  |  |  |  |
| - No. observations | - | - | - | - | - |  |
| - Male | 438 [13%] | 2353 [8%] | 44 [7%] | 6189 [12%] | 280 [9%] | <0.001 |
| - Female | 2990 [87%] | 26192 [92%] | 571 [93%] | 46650 [88%] | 2759 [91%] |  |
| Marital status (n [%]) |  |  |  |  |  |  |
| - No. observations | 3069 [90%] | - | - | - | - |  |
| - Living together | 2026 [66%] | 22707 [80%] | 469 [76%] | 41957 [79%] | 2282 [75%] | <0.001 |
| - Not living together | 22 [1%] | 437 [2%] | 13 [2%] | 1701 [3%] | 131 [4%] |  |
| - Not in a relationship | 1021 [33%] | 5401 [19%] | 133 [22%] | 9181 [17%] | 626 [21%] |  |
| Employment status (n [%]) |  |  |  |  |  |  |
| - No. observations | 783 [23%] | - | - | - | - |  |
| - Active | 472 [60%] | 17701 [62%] | 485 [79%] | 44041 [83%] | 2562 [84%] | 0.18 |
| - Unemployed/Inactive | 311 [40%] | 10844 [38%] | 130 [21%] | 8798 [17%] | 477 [16%] |  |
| Educational level (n [%]) |  |  |  |  |  |  |
| - No. observations | 2795 [82%] | - | - | - | - |  |
| - Elementary school | 1137 [41%] | 4226 [15%] | 46 [7%] | 3306 [6%] | 165 [5%] | 0.07 |
| - At least secondary school | 1658 [59%] | 24319 [85%] | 569 [93%] | 49533 [94%] | 2874 [95%] |  |
| Self-reported call duration (n [%]) |  |  |  |  |  |  |
| - No. observations | - | - | - | - | - |  |
| < 5 min/week | 338 [10%] | - | - | 7757 [15%] | 378 [12%] | <0.001 |
| 5-29 min/week | 726 [21%] | - | - | 23867 [45%] | 1555 [51%] |  |
| 30-59 min/week | 186 [5%] | - | - | 9811 [19%] | 642 [21%] |  |
| 1-3 hours/week | 123 [4%] | - | - | 7751 [15%] | 359 [12%] |  |
| 4-6 hours/week | 29 [1%] | - | - | 2219 [4%] | 75 [2%] |  |
| >6 hours/week | 2026 [59%] | - | - | 1434 [3%] | 30 [1%] |  |
| Recorded call duration |  |  |  |  |  |  |
| - No. observations | 55 [2%] | - | - | - | - |  |
| - Minutes/week (GM [GSD]) | 20.0 [3.9] | - | 10.3 [3.2] | - | 23.4 [2.6] |  |

|  | **Sweden** | | | | |  |
| --- | --- | --- | --- | --- | --- | --- |
|  | Covariate data missing | Self-reported & operator-recorded data missing | Non-users or self-reported data missing | Operator-recorded data missing | Complete data available | P |
| Number of participants (n) | 2215 | 4829 | 130 | 18623 | 24881 |  |
| Age |  |  |  |  |  |  |
| - No. observations | 2037 [92%] | - | - | - | - |  |
| - Years (mean [sd]) | 47.5 [14.2] | 50.0 [14.3] | 48.2 [13.1] | 44.8 [13.5] | 43.3 [13.4] | <0.001 |
| Sex (n [%]) |  |  |  |  |  |  |
| - No. observations | 2037 [92%] | - | - | - | - |  |
| - Male | 774 [38%] | 1979 [41%] | 58 [45%] | 9497 [51%] | 11352 [46%] | <0.001 |
| - Female | 1263 [62%] | 2850 [59%] | 72 [55%] | 9126 [49%] | 13529 [54%] |  |
| Marital status (n [%]) |  |  |  |  |  |  |
| - No. observations | 1262 [57%] | - | - | - | - |  |
| - Living together | 757 [60%] | 3187 [66%] | 89 [69%] | 12477 [67%] | 16784 [67%] | <0.001 |
| - Not living together | 139 [11%] | 241 [5%] | 13 [10%] | 1862 [10%] | 2768 [11%] |  |
| - Not in a relationship | 366 [29%] | 1401 [29%] | 28 [21%] | 4284 [23%] | 5329 [21%] |  |
| Employment status (n [%]) |  |  |  |  |  |  |
| - No. observations | 376 [17%] | - | - | - | - |  |
| - Active | 192 [51%] | 2945 [61%] | 89 [69%] | 13967 [75%] | 18033 [72%] | <0.001 |
| - Unemployed/Inactive | 184 [49%] | 1884 [39%] | 41 [31%] | 4656 [25%] | 6848 [28%] |  |
| Educational level (n [%]) |  |  |  |  |  |  |
| - No. observations | 1661 [75%] | - | - | - | - |  |
| - Elementary school | 415 [25%] | 1352 [28%] | 23 [18%] | 2897 [16%] | 3331 [13%] | <0.001 |
| - At least secondary school | 1246 [75%] | 3477 [72%] | 107 [82%] | 15214 [84%] | 21550 [87%] |  |
| Self-reported call duration (n [%]) |  |  |  |  |  |  |
| - No. observations | - | - | - | - | - |  |
| < 5 min/week | 133 [6%] | - | - | 1303 [7%] | 1021 [4%] | <0.001 |
| 5-29min/week | 488 [22%] | - | - | 5028 [27%] | 7023 [28%] |  |
| 30-59 min/week | 354 [16%] | - | - | 3352 [18%] | 4894 [20%] |  |
| 1-3 hours/week | 453 [20%] | - | - | 4469 [24%] | 6647 [27%] |  |
| 4-6 hours/week | 188 [8%] | - | - | 2420 [13%] | 3075 [12%] |  |
| >6 hours/week | 599 [27%] | - | - | 2051 [11%] | 2221 [9%] |  |
| Recorded call duration |  |  |  |  |  |  |
| - No. observations | 863 [39%] | - | - | - | - |  |
| - Minutes/week (GM [GSD]) | 83.2 [3.9] | - | 57.8 [5.1] | - | 78.3 [3.9] |  |

|  | **United Kingdom** | | | | |  |
| --- | --- | --- | --- | --- | --- | --- |
|  | Covariate data missing | Self-reported & operator-recorded data missing | Non-users or self-reported data missing | Operator-recorded data missing | Complete data available | P |
| Number of participants (n) | 12008 | 6336 | 0 | 23479 | 56862 |  |
| Age |  |  |  |  |  |  |
| - No. observations | - | - | - | - | - |  |
| - Years (mean [sd]) | 43.6 [14.8] | 50.7 [17.2] | - | 44.6 [14.7] | 45.1 [14.7] | <0.001 |
| Sex (n [%]) |  |  |  |  |  |  |
| - No. observations | - | - | - | - | - |  |
| - Male | 5749 [48%] | 2799 [44%] | - | 12257 [52.2%] | 26044 [45.8%] | <0.001 |
| - Female | 6259 [52%] | 3537 [56%] | - | 11222 [47.8%] | 30818 [54.2%] |  |
| Marital status (n [%]) |  |  |  |  |  |  |
| - No. observations | 2739 [23%] | - | - | - | - |  |
| - Living together | 1965 [72%] | 4223 [67%] | - | 16254 [69.2%] | 37304 [65.6%] | <0.001 |
| - Not living together | 241 [9%] | 416 [7%] | - | 2731 [11.6%] | 7445 [13.1%] |  |
| - Not in a relationship | 533 [19%] | 1697 [27%] | - | 4494 [19.1%] | 12113 [21.3%] |  |
| Employment status (n [%]) |  |  |  |  |  |  |
| - No. observations | 2805 [23%] | - | - | - | - |  |
| - Active | 1725 [61%] | 3103 [49%] | - | 16680 [71%] | 39854 [70.1%] |  |
| - Unemployed/Inactive | 1080 [39%] | 3233 [51%] | - | 6799 [29%] | 17008 [29.9%] | 0.007 |
| Educational level (n [%]) |  |  |  |  |  |  |
| - No. observations | 498 [4%] | - | - | - | - |  |
| - Elementary school | 65 [13%] | 711 [11%] | - | 2702 [11.5%] | 6166 [10.8%] |  |
| - At least secondary school | 433 [87%] | 5625 [89%] | - | 20777 [88.5%] | 50696 [89.2%] | 0.007 |
| Self-reported call duration (n [%]) |  |  |  |  |  |  |
| - No. observations | 9483 [79%] | - | - | - | - |  |
| < 5 min/week | 374 [4%] | - | - | 1391 [5.9%] | 2471 [4.3%] | <0.001 |
| 5-29min/week | 2271 [24%] | - | - | 6596 [28.1%] | 15651 [27.5%] |  |
| 30-59 min/week | 1967 [21%] | - | - | 5040 [21.5%] | 12754 [22.4%] |  |
| 1-3 hours/week | 2669 [28%] | - | - | 5987 [25.5%] | 15610 [27.5%] |  |
| 4-6 hours/week | 1204 [13%] | - | - | 2450 [10.4%] | 6030 [10.6%] |  |
| >6 hours/week | 998 [11%] | - | - | 2015 [8.6%] | 4346 [7.6%] |  |
| Recorded call duration |  |  |  |  |  |  |
| - No. observations | 5346 [45%] | - | - | - | - |  |
| - Minutes/week (GM [GSD]) | 40.6 [4.0] | - | - | - | 46.5 [3.9] |  |

**Table A3A.** Point estimates, percentiles, squared bias, variance and MSE for the different approaches based on the results from 1,000 simulations (100 simulations for the Inverse RC approach). Simulations used data from 50% of the participants with complete information on both self-reported and operator-recorded data as the training set and the remainder as test set. The outcome was simulated using a slope coefficient (β) of 0.001 (i.e. assuming an Odds Ratio of exp(0.02)=1.02 for each additional 20 minutes call-time per week) and with a balanced ratio of cases:non-cases. Bootstrapping was used to estimate 95%CIs for each statistic. The training set was used to fit the health model for the CC analysis, to fit the first stage (exposure) models for the RC approaches, and to fit the MI model. All second stage (health) models for the RC approaches were fitted to the test data only and results were precision-weighted with those from the CC approach before further analyses. The (non-full data) model that achieved lowest MSE and all models for which the estimated MSE fell within the 95%CI for that lowest MSE are highlighted in grey.

| **Country** | **Model** | **β (*1000)  [95%CI]** | **Percentiles^$^ (2.5%,97.5%)** | **Bias^2^  [95%CI]** | **Variance  [95%CI]** | **MSE^#^  [95%CI]** |
| --- | --- | --- | --- | --- | --- | --- |
| Denmark | Full data | 1.02 | (0.33, 1.74) | 0.00 | 0.19 | 0.19 |
|  |  | [0.99, 1.04] |  | [0.00, 0.00] | [0.17, 0.21] | [0.17, 0.21] |
|  | Simple RC | 1.01 | (0.20, 1.82) | 0.00 | 0.26 | 0.26 |
|  |  | [0.98, 1.04] |  | [0.00, 0.00] | [0.24, 0.28] | [0.24, 0.28] |
|  | GAMLSS RC | 0.94 | (0.18, 1.74) | 0.00 | 0.23 | 0.23 |
|  |  | [0.91, 0.97] |  | [0.00, 0.01] | [0.21, 0.24] | [0.21, 0.25] |
|  | Direct RC | 1.00 | (0.21, 1.83) | 0.00 | 0.25 | 0.25 |
|  |  | [0.97, 1.04] |  | [0.00, 0.00] | [0.23, 0.27] | [0.23, 0.27] |
|  | Inverse RC | 0.93 | (0.18, 1.82) | 0.00 | 0.24 | 0.24 |
|  |  | [0.84, 1.03] |  | [0.00, 0.03] | [0.18, 0.32] | [0.18, 0.32] |
|  | CC | 1.03 | (0.06, 1.99) | 0.00 | 0.35 | 0.35 |
|  |  | [0.99, 1.07] |  | [0.00, 0.00] | [0.32, 0.38] | [0.32, 0.38] |
|  | MI | 1.00 | (0.17, 1.84) | 0.00 | 0.28 | 0.28 |
|  |  | [0.96, 1.03] |  | [0.00, 0.00] | [0.26, 0.31] | [0.26, 0.31] |
| Finland | Full data | 0.99 | (0.54, 1.45) | 0.00 | 0.08 | 0.08 |
|  |  | [0.97, 1.00] |  | [0.00, 0.00] | [0.07, 0.09] | [0.07, 0.09] |
|  | Simple RC | 0.99 | (0.41, 1.59) | 0.00 | 0.13 | 0.13 |
|  |  | [0.97, 1.01] |  | [0.00, 0.00] | [0.11, 0.14] | [0.12, 0.14] |
|  | GAMLSS RC | 0.87 | (0.37, 1.40) | 0.02 | 0.10 | 0.12 |
|  |  | [0.85, 0.89] |  | [0.01, 0.02] | [0.09, 0.11] | [0.11, 0.13] |
|  | Direct RC | 0.99 | (0.44, 1.59) | 0.00 | 0.12 | 0.12 |
|  |  | [0.97, 1.01] |  | [0.00, 0.00] | [0.11, 0.13] | [0.11, 0.13] |
|  | Inverse RC | 1.04 | (0.49, 1.61) | 0.00 | 0.13 | 0.13 |
|  |  | [0.97, 1.11] |  | [0.00, 0.01] | [0.11, 0.17] | [0.11, 0.17] |
|  | CC | 0.99 | (0.36, 1.66) | 0.00 | 0.16 | 0.16 |
|  |  | [0.97, 1.02] |  | [0.00, 0.00] | [0.14, 0.17] | [0.14, 0.17] |
|  | MI | 1.02 | (0.41, 1.69) | 0.00 | 0.15 | 0.15 |
|  |  | [0.99, 1.04] |  | [0.00, 0.00] | [0.14, 0.17] | [0.14, 0.17] |
| Netherlands | Full data | 0.95 | (-1.83, 3.44) | 0.00 | 2.46 | 2.47 |
|  |  | [0.86, 1.05] |  | [0.00, 0.02] | [2.26, 2.68] | [2.26, 2.70] |
|  | Simple RC | 0.94 | (-2.30, 4.09) | 0.00 | 3.76 | 3.76 |
|  |  | [0.82, 1.06] |  | [0.00, 0.03] | [3.44, 4.12] | [3.44, 4.12] |
|  | GAMLSS RC | 0.91 | (-2.09, 3.99) | 0.01 | 3.45 | 3.46 |
|  |  | [0.80, 1.03] |  | [0.00, 0.04] | [3.18, 3.76] | [3.18, 3.76] |
|  | Direct RC | 0.89 | (-2.12, 3.99) | 0.01 | 3.37 | 3.38 |
|  |  | [0.76, 0.99] |  | [0.00, 0.06] | [3.09, 3.66] | [3.10, 3.67] |
|  | Inverse RC | 1.00 | (-2.29, 4.15) | 0.00 | 3.46 | 3.46 |
|  |  | [0.63, 1.39] |  | [0.00, 0.00] | [2.63, 4.59] | [2.59, 4.52] |
|  | CC | 0.94 | (-2.59, 4.62) | 0.00 | 5.09 | 5.09 |
|  |  | [0.80, 1.08] |  | [0.00, 0.03] | [4.65, 5.66] | [4.66, 5.67] |
|  | MI | 0.96 | (-2.45, 4.48) | 0.00 | 4.47 | 4.47 |
|  |  | [0.83, 1.09] |  | [0.00, 0.01] | [4.09, 4.84] | [4.09, 4.86] |
| Sweden | Full data | 1.00 | (0.78, 1.21) | 0.00 | 0.02 | 0.02 |
|  |  | [0.99, 1.01] |  | [0.00, 0.00] | [0.02, 0.02] | [0.02, 0.02] |
|  | Simple RC | 1.00 | (0.74, 1.28) | 0.00 | 0.03 | 0.03 |
|  |  | [0.99, 1.01] |  | [0.00, 0.00] | [0.02, 0.03] | [0.02, 0.03] |
|  | GAMLSS RC | 0.86 | (0.64, 1.09) | 0.02 | 0.02 | 0.04 |
|  |  | [0.85, 0.87] |  | [0.02, 0.02] | [0.02, 0.02] | [0.04, 0.04] |
|  | Direct RC | 1.00 | (0.74, 1.27) | 0.00 | 0.03 | 0.03 |
|  |  | [0.99, 1.01] |  | [0.00, 0.00] | [0.02, 0.03] | [0.02, 0.03] |
|  | Inverse RC | 1.03 | (0.79, 1.25) | 0.00 | 0.02 | 0.02 |
|  |  | [1.00, 1.06] |  | [0.00, 0.00] | [0.02, 0.03] | [0.02, 0.03] |
|  | CC | 1.00 | (0.68, 1.32) | 0.00 | 0.04 | 0.04 |
|  |  | [0.99, 1.01] |  | [0.00, 0.00] | [0.03, 0.04] | [0.03, 0.04] |
|  | MI | 1.04 | (0.73, 1.34) | 0.00 | 0.03 | 0.04 |
|  |  | [1.02, 1.05] |  | [0.00, 0.00] | [0.03, 0.04] | [0.03, 0.04] |
| United Kingdom | Full data | 1.00 | (0.81, 1.18) | 0.00 | 0.01 | 0.01 |
|  |  | [0.99, 1.00] |  | [0.00, 0.00] | [0.01, 0.01] | [0.01, 0.01] |
|  | Simple RC | 0.99 | (0.77, 1.22) | 0.00 | 0.02 | 0.02 |
|  |  | [0.98, 1.00] |  | [0.00, 0.00] | [0.02, 0.02] | [0.02, 0.02] |
|  | GAMLSS RC | 0.95 | (0.74, 1.17) | 0.00 | 0.02 | 0.02 |
|  |  | [0.95, 0.96] |  | [0.00, 0.00] | [0.02, 0.02] | [0.02, 0.02] |
|  | Direct RC | 0.99 | (0.77, 1.22) | 0.00 | 0.02 | 0.02 |
|  |  | [0.98, 1.00] |  | [0.00, 0.00] | [0.02, 0.02] | [0.02, 0.02] |
|  | Inverse RC | 1.05 | (0.69, 1.41) | 0.00 | 0.05 | 0.05 |
|  |  | [1.01, 1.09] |  | [0.00, 0.01] | [0.04, 0.06] | [0.04, 0.07] |
|  | CC | 1.00 | (0.74, 1.26) | 0.00 | 0.02 | 0.02 |
|  |  | [0.99, 1.00] |  | [0.00, 0.00] | [0.02, 0.03] | [0.02, 0.03] |
|  | MI | 1.00 | (0.79, 1.23) | 0.00 | 0.02 | 0.02 |
|  |  | [0.98, 1.03] |  | [0.00, 0.00] | [0.01, 0.02] | [0.01, 0.02] |

CC, complete case; CI, confidence interval; MI, multiple imputation; RC, regression calibration

**Table A3B.** Point estimates, squared bias, variance and MSE for the different approaches based on the results from 1,000 simulations (100 simulations for the Inverse RC approach). Simulations used data from 25% of the participants with complete information on both self-reported and operator-recorded data as the training set and the remainder as test set. The outcome was simulated using a slope coefficient (β) of 0.005 (i.e. assuming an Odds Ratio of exp(0.1)=1.11 for each additional 20 minutes call-time per week) and with a balanced ratio of cases:non-cases. Bootstrapping was used to estimate 95%CIs for each statistic. The training set was used to fit the health model for the CC analysis, to fit the first stage (exposure) models for the RC approaches, and to fit the multiple imputation model. All second stage (health) models for the RC approaches were fitted to the test data only and results were precision-weighted with those from the CC approach before further analyses. The (non-full data) model that achieved lowest MSE and all models for which the estimated MSE fell within the 95%CI for that lowest MSE are highlighted in grey.

| **Country** | **Model** | | **β (*1000)  [95%CI]** | **Percentiles^$^ (2.5%,97.5%)** | **Bias^2^  [95%CI]** | **Variance  [95%CI]** | **MSE^#^  [95%CI]** |
| --- | --- | --- | --- | --- | --- | --- | --- |
| Denmark | | Full data | 5.02 | (4.16, 5.90) | 0.00 | 0.28 | 0.29 |
|  | |  | [4.98, 5.05] |  | [0.00, 0.00] | [0.26, 0.31] | [0.26, 0.31] |
|  | | Simple RC | 4.68 | (3.61, 5.82) | 0.10 | 0.46 | 0.56 |
|  | |  | [4.64, 4.72] |  | [0.08, 0.13] | [0.42, 0.50] | [0.52, 0.61] |
|  | | GAMLSS RC | 4.05 | (2.99, 5.17) | 0.91 | 0.43 | 1.34 |
|  | |  | [4.00, 4.09] |  | [0.83, 0.99] | [0.40, 0.48] | [1.26, 1.43] |
|  | | Direct RC | 4.60 | (3.57, 5.73) | 0.16 | 0.43 | 0.59 |
|  | |  | [4.56, 4.64] |  | [0.13, 0.19] | [0.40, 0.47] | [0.54, 0.64] |
|  | | Inverse RC | 4.40 | (3.47, 5.37) | 0.36 | 0.36 | 0.71 |
|  | |  | [4.28, 4.52] |  | [0.23, 0.52] | [0.27, 0.48] | [0.57, 0.89] |
|  | | CC | 5.05 | (3.29, 6.84) | 0.00 | 1.15 | 1.15 |
|  | |  | [4.98, 5.12] |  | [0.00, 0.01] | [1.05, 1.25] | [1.05, 1.25] |
|  | | MI | 4.69 | (3.37, 6.12) | 0.10 | 0.70 | 0.80 |
|  | |  | [4.64, 4.75] |  | [0.06, 0.13] | [0.64, 0.76] | [0.73, 0.87] |
| Finland | | Full data | 5.00 | (4.45, 5.55) | 0.00 | 0.11 | 0.11 |
|  | |  | [4.98, 5.02] |  | [0.00, 0.00] | [0.11, 0.13] | [0.11, 0.13] |
|  | | Simple RC | 4.78 | (4.00, 5.58) | 0.05 | 0.24 | 0.29 |
|  | |  | [4.75, 4.81] |  | [0.04, 0.06] | [0.22, 0.26] | [0.26, 0.31] |
|  | | GAMLSS RC | 3.58 | (2.76, 4.40) | 2.01 | 0.26 | 2.26 |
|  | |  | [3.55, 3.62] |  | [1.92, 2.10] | [0.24, 0.28] | [2.17, 2.36] |
|  | | Direct RC | 4.76 | (4.02, 5.52) | 0.06 | 0.23 | 0.28 |
|  | |  | [4.73, 4.79] |  | [0.04, 0.07] | [0.21, 0.25] | [0.26, 0.31] |
|  | | Inverse RC | 5.05 | (4.22, 5.91) | 0.00 | 0.27 | 0.27 |
|  | |  | [4.95, 5.16] |  | [0.00, 0.02] | [0.20, 0.40] | [0.20, 0.41] |
|  | | CC | 5.04 | (3.95, 6.20) | 0.00 | 0.47 | 0.47 |
|  | |  | [5.00, 5.08] |  | [0.00, 0.01] | [0.43, 0.51] | [0.43, 0.52] |
|  | | MI | 5.09 | (4.00, 6.27) | 0.01 | 0.47 | 0.47 |
|  | |  | [5.05, 5.13] |  | [0.00, 0.02] | [0.43, 0.51] | [0.43, 0.52] |
| Netherlands | | Full data | 5.10 | (2.38, 7.97) | 0.01 | 2.94 | 2.95 |
|  | |  | [4.99, 5.20] |  | [0.00, 0.04] | [2.70, 3.21] | [2.70, 3.23] |
|  | | Simple RC | 5.05 | (1.07, 9.07) | 0.00 | 5.79 | 5.79 |
|  | |  | [4.90, 5.19] |  | [0.00, 0.02] | [5.32, 6.31] | [5.32, 6.32] |
|  | | GAMLSS RC | 4.57 | (0.83, 8.29) | 0.19 | 5.09 | 5.28 |
|  | |  | [4.44, 4.70] |  | [0.09, 0.32] | [4.68, 5.60] | [4.86, 5.80] |
|  | | Direct RC | 3.68 | (0.57, 6.74) | 1.74 | 3.75 | 5.50 |
|  | |  | [3.56, 3.80] |  | [1.45, 2.07] | [3.45, 4.14] | [5.09, 5.96] |
|  | | Inverse RC | 4.46 | (0.64, 8.74) | 0.30 | 6.40 | 6.69 |
|  | |  | [3.98, 5.04] |  | [0.00, 1.03] | [4.99, 8.20] | [5.24, 8.44] |
|  | | CC | 5.18 | (-0.42, 11.36) | 0.03 | 12.76 | 12.80 |
|  | |  | [4.99, 5.42] |  | [0.00, 0.17] | [11.71, 13.90] | [11.75, 14.02] |
|  | | MI | 5.14 | (0.31, 10.22) | 0.02 | 9.30 | 9.32 |
|  | |  | [4.95, 5.33] |  | [0.00, 0.11] | [8.53, 10.25] | [8.56, 10.31] |
| Sweden | | Full data | 5.00 | (4.74, 5.28) | 0.00 | 0.03 | 0.03 |
|  | |  | [4.99, 5.01] |  | [0.00, 0.00] | [0.03, 0.03] | [0.03, 0.03] |
|  | | Simple RC | 4.71 | (4.36, 5.10) | 0.08 | 0.05 | 0.13 |
|  | |  | [4.70, 4.73] |  | [0.07, 0.09] | [0.05, 0.06] | [0.13, 0.14] |
|  | | GAMLSS RC | 3.59 | (3.17, 4.00) | 1.99 | 0.06 | 2.05 |
|  | |  | [3.57, 3.60] |  | [1.95, 2.03] | [0.06, 0.07] | [2.01, 2.10] |
|  | | Direct RC | 4.69 | (4.34, 5.08) | 0.09 | 0.05 | 0.14 |
|  | |  | [4.68, 4.71] |  | [0.09, 0.10] | [0.05, 0.06] | [0.14, 0.15] |
|  | | Inverse RC | 5.12 | (4.70, 5.58) | 0.01 | 0.08 | 0.09 |
|  | |  | [5.06, 5.17] |  | [0.00, 0.03] | [0.06, 0.11] | [0.07, 0.13] |
|  | | CC | 5.01 | (4.48, 5.54) | 0.00 | 0.11 | 0.11 |
|  | |  | [4.99, 5.03] |  | [0.00, 0.00] | [0.10, 0.12] | [0.10, 0.12] |
|  | | MI | 5.25 | (4.70, 5.84) | 0.06 | 0.13 | 0.19 |
|  | |  | [5.23, 5.27] |  | [0.05, 0.07] | [0.11, 0.14] | [0.17, 0.20] |
| United | | Full data | 5.00 | (4.76, 5.23) | 0.00 | 0.02 | 0.02 |
| Kingdom | |  | [4.99, 5.01] |  | [0.00, 0.00] | [0.02, 0.02] | [0.02, 0.02] |
|  | | Simple RC | 4.67 | (4.38, 4.97) | 0.11 | 0.03 | 0.14 |
|  | |  | [4.66, 4.68] |  | [0.10, 0.11] | [0.03, 0.03] | [0.13, 0.15] |
|  | | GAMLSS RC | 4.28 | (3.99, 4.55) | 0.52 | 0.03 | 0.55 |
|  | |  | [4.27, 4.29] |  | [0.51, 0.54] | [0.03, 0.03] | [0.54, 0.57] |
|  | | Direct RC | 4.64 | (4.36, 4.93) | 0.13 | 0.03 | 0.16 |
|  | |  | [4.63, 4.66] |  | [0.12, 0.13] | [0.03, 0.03] | [0.15, 0.17] |
|  | | Inverse RC | 4.81 | (4.48, 5.16) | 0.03 | 0.05 | 0.09 |
|  | |  | [4.76, 4.86] |  | [0.02, 0.06] | [0.04, 0.08] | [0.07, 0.11] |
|  | | CC | 4.99 | (4.54, 5.45) | 0.00 | 0.07 | 0.07 |
|  | |  | [4.97, 5.00] |  | [0.00, 0.00] | [0.07, 0.08] | [0.07, 0.08] |
|  | | MI | 4.88 | (4.59, 5.16) | 0.01 | 0.04 | 0.05 |
|  | |  | [4.85, 4.93] |  | [0.01, 0.02] | [0.03, 0.05] | [0.04, 0.07] |

CC, complete case; CI, confidence interval; MI, multiple imputation; RC, regression calibration.

**Table A4A.** Coverage of 95%CI and ratio of slope estimate (β) over its standard error (SE) as a proxy for efficiency for the different approaches based on the results from 1,000 simulations (100 simulations for the Inverse RC approach). Simulations used data from 50% of the participants with complete information on both self-reported and operator-recorded data as the training set and the remainder as test set. The outcome was simulated using a slope coefficient (β) of 0.001 (i.e. assuming an Odds Ratio of exp(0.02)=1.02 for each additional 20 minutes call-time per week) and with a balanced ratio of cases:non-cases. Bootstrapping was used to estimate 95%CIs for each statistic.

| **Country** | **Model** | **Coverage  [95%CI]** | **β/SE  [95%CI]** | **Percentiles^$^ (2.5%, 97.5%)** |
| --- | --- | --- | --- | --- |
| Denmark | Full data | 95% | 2.38 | (0.79, 3.94) |
|  |  | [93%, 96%] | [2.31, 2.43] |  |
|  | Simple RC | 96% | 1.95 | (0.39, 3.48) |
|  |  | [95%, 97%] | [1.89, 2.01] |  |
|  | GAMLSS RC | 96% | 1.95 | (0.40, 3.51) |
|  |  | [94%, 97%] | [1.89, 2.02] |  |
|  | Direct RC | 96% | 1.97 | (0.41, 3.49) |
|  |  | [95%, 97%] | [1.90, 2.03] |  |
|  | Inverse RC | 94% | 1.84 | (0.34, 3.56) |
|  |  | [86%, 97%] | [1.65, 2.03] |  |
|  | CC | 97% | 1.68 | (0.11, 3.13) |
|  |  | [95%, 98%] | [1.62, 1.74] |  |
|  | MI | 96% | 1.77 | (0.35, 3.17) |
|  |  | [95%, 97%] | [1.71, 1.83] |  |
| Finland | Full data | 95% | 3.52 | (1.99, 5.05) |
|  |  | [93%, 96%] | [3.47, 3.58] |  |
|  | Simple RC | 95% | 2.79 | (1.23, 4.41) |
|  |  | [93%, 96%] | [2.73, 2.85] |  |
|  | GAMLSS RC | 93% | 2.76 | (1.18, 4.36) |
|  |  | [91%, 94%] | [2.70, 2.82] |  |
|  | Direct RC | 95% | 2.83 | (1.28, 4.43) |
|  |  | [93%, 96%] | [2.77, 2.89] |  |
|  | Inverse RC | 96% | 2.85 | (1.39, 4.28) |
|  |  | [89%, 98%] | [2.66, 3.03] |  |
|  | CC | 95% | 2.50 | (0.93, 4.07) |
|  |  | [93%, 96%] | [2.44, 2.56] |  |
|  | MI | 95% | 2.56 | (1.08, 4.15) |
|  |  | [94%, 96%] | [2.50, 2.62] |  |
| Netherlands | Full data | 96% | 0.58 | (-1.11, 2.07) |
|  |  | [95%, 97%] | [0.52, 0.63] |  |
|  | Simple RC | 96% | 0.46 | (-1.11, 2.01) |
|  |  | [95%, 97%] | [0.40, 0.52] |  |
|  | GAMLSS RC | 96% | 0.47 | (-1.08, 1.99) |
|  |  | [94%, 97%] | [0.41, 0.53] |  |
|  | Direct RC | 96% | 0.46 | (-1.09, 2.03) |
|  |  | [94%, 97%] | [0.40, 0.52] |  |
|  | Inverse RC | 96% | 0.49 | (-1.22, 1.92) |
|  |  | [88%, 98%] | [0.31, 0.68] |  |
|  | CC | 96% | 0.40 | (-1.11, 1.95) |
|  |  | [94%, 97%] | [0.34, 0.46] |  |
|  | MI | 97% | 0.42 | (-1.07, 1.98) |
|  |  | [96%, 98%] | [0.36, 0.48] |  |
| Sweden | Full data | 95% | 7.67 | (6.07, 9.18) |
|  |  | [93%, 96%] | [7.61, 7.72] |  |
|  | Simple RC | 95% | 6.34 | (4.73, 8.01) |
|  |  | [93%, 96%] | [6.28, 6.39] |  |
|  | GAMLSS RC | 81% | 6.25 | (4.65, 7.90) |
|  |  | [79%, 84%] | [6.19, 6.31] |  |
|  | Direct RC | 94% | 6.37 | (4.72, 8.02) |
|  |  | [93%, 95%] | [6.31, 6.43] |  |
|  | Inverse RC | 95% | 6.34 | (5.01, 7.84) |
|  |  | [86%, 98%] | [6.16, 6.52] |  |
|  | CC | 94% | 5.44 | (3.73, 7.06) |
|  |  | [92%, 95%] | [5.38, 5.49] |  |
|  | MI | 94% | 5.60 | (3.80, 7.54) |
|  |  | [92%, 95%] | [5.53, 5.67] |  |
| United Kingdom | Full data | 95% | 9.10 | (7.52, 10.64) |
|  |  | [93%, 96%] | [9.04, 9.16] |  |
|  | Simple RC | 95% | 7.38 | (5.79, 8.97) |
|  |  | [93%, 96%] | [7.32, 7.44] |  |
|  | GAMLSS RC | 93% | 7.41 | (5.80, 9.04) |
|  |  | [91%, 94%] | [7.34, 7.46] |  |
|  | Direct RC | 95% | 7.41 | (5.83, 9.02) |
|  |  | [93%, 96%] | [7.35, 7.47] |  |
|  | Inverse RC | 94% | 5.13 | (3.38, 6.87) |
|  |  | [86%, 97%] | [4.92, 5.32] |  |
|  | CC | 95% | 6.41 | (4.88, 7.94) |
|  |  | [93%, 96%] | [6.36, 6.47] |  |
|  | MI | 96% | 7.35 | (5.83, 8.92) |
|  |  | [89%, 98%] | [7.16, 7.55] |  |

CC, complete case; CI, confidence interval; MI, multiple imputation; RC, regression calibration.

**Table A4B.** Coverage of 95%CI and ratio of slope estimate (β) over its standard error (SE) as a proxy for efficiency for the different approaches based on the results from 1,000 simulations (100 simulations for the Inverse RC approach). Simulations used data from 25% of the participants with complete information on both self-reported and operator-recorded data as the training set and the remainder as test set. The outcome was simulated using a slope coefficient (β) of 0.005 (i.e. assuming an Odds Ratio of exp(0.1)=1.11 for each additional 20 minutes call-time per week) and with a balanced ratio of cases:non-cases. Bootstrapping was used to estimate 95%CIs for each statistic.

| **Country** | **Model** | **Coverage  [95%CI]** | **β/SE  [95%CI]** | **Percentiles^$^ (2.5%, 97.5%)** |
| --- | --- | --- | --- | --- |
| Denmark | Full data | 96% | 9.22 | (8.06, 10.36) |
|  |  | [94%, 97%] | [9.17, 9.26] |  |
|  | Simple RC | 89% | 7.25 | (5.82, 8.64) |
|  |  | [87%, 91%] | [7.19, 7.29] |  |
|  | GAMLSS RC | 57% | 7.18 | (5.78, 8.65) |
|  |  | [54%, 60%] | [7.13, 7.24] |  |
|  | Direct RC | 86% | 7.34 | (5.94, 8.73) |
|  |  | [84%, 88%] | [7.28, 7.39] |  |
|  | Inverse RC | 82% | 7.21 | (5.98, 8.67) |
|  |  | [72%, 88%] | [7.07, 7.35] |  |
|  | CC | 96% | 4.60 | (3.32, 5.73) |
|  |  | [94%, 97%] | [4.56, 4.65] |  |
|  | MI | 96% | 4.64 | (3.15, 6.44) |
|  |  | [94%, 97%] | [4.58, 4.71] |  |
| Finland | Full data | 96% | 14.80 | (13.49, 16.05) |
|  |  | [94%, 97%] | [14.75, 14.85] |  |
|  | Simple RC | 90% | 10.24 | (8.85, 11.60) |
|  |  | [88%, 92%] | [10.19, 10.30] |  |
|  | GAMLSS RC | 10% | 9.96 | (8.43, 11.43) |
|  |  | [8%, 12%] | [9.90, 10.02] |  |
|  | Direct RC | 90% | 10.42 | (9.01, 11.85) |
|  |  | [88%, 91%] | [10.37, 10.48] |  |
|  | Inverse RC | 95% | 10.13 | (8.73, 11.22) |
|  |  | [89%, 98%] | [9.97, 10.30] |  |
|  | CC | 96% | 7.42 | (6.10, 8.70) |
|  |  | [94%, 97%] | [7.37, 7.47] |  |
|  | MI | 96% | 7.14 | (4.92, 9.88) |
|  |  | [94%, 97%] | [7.05, 7.24] |  |
| Netherlands | Full data | 95% | 2.99 | (1.44, 4.53) |
|  |  | [94%, 96%] | [2.93, 3.05] |  |
|  | Simple RC | 95% | 2.10 | (0.47, 3.63) |
|  |  | [94%, 96%] | [2.04, 2.16] |  |
|  | GAMLSS RC | 93% | 2.06 | (0.39, 3.64) |
|  |  | [91%, 94%] | [2.01, 2.12] |  |
|  | Direct RC | 85% | 1.97 | (0.29, 3.53) |
|  |  | [82%, 87%] | [1.91, 2.03] |  |
|  | Inverse RC | 95% | 1.89 | (0.27, 3.60) |
|  |  | [87%, 97%] | [1.70, 2.12] |  |
|  | CC | 95% | 1.48 | (-0.12, 3.04) |
|  |  | [93%, 96%] | [1.43, 1.55] |  |
|  | MI | 96% | 1.58 | (0.09, 2.99) |
|  |  | [94%, 97%] | [1.52, 1.64] |  |
| Sweden | Full data | 95% | 30.56 | (29.43, 31.76) |
|  |  | [93%, 96%] | [30.51, 30.61] |  |
|  | Simple RC | 65% | 23.75 | (22.38, 25.23) |
|  |  | [62%, 68%] | [23.69, 23.81] |  |
|  | GAMLSS RC | 0% | 23.42 | (21.88, 24.99) |
|  |  | [0%, 0%] | [23.37, 23.48] |  |
|  | Direct RC | 61% | 23.93 | (22.50, 25.43) |
|  |  | [58%, 64%] | [23.88, 23.99] |  |
|  | Inverse RC | 83% | 23.89 | (22.42, 25.52) |
|  |  | [73%, 88%] | [23.69, 24.12] |  |
|  | CC | 95% | 15.29 | (14.06, 16.48) |
|  |  | [94%, 96%] | [15.24, 15.33] |  |
|  | MI | 90% | 14.34 | (10.18, 19.70) |
|  |  | [87%, 91%] | [14.16, 14.53] |  |
| United Kingdom | Full data | 94% | 35.73 | (34.46, 36.97) |
|  |  | [93%, 96%] | [35.68, 35.77] |  |
|  | Simple RC | 54% | 26.82 | (25.48, 28.15) |
|  |  | [50%, 56%] | [26.77, 26.88] |  |
|  | GAMLSS RC | 1% | 27.06 | (25.68, 28.45) |
|  |  | [1%, 2%] | [27.01, 27.11] |  |
|  | Direct RC | 47% | 27.09 | (25.74, 28.47) |
|  |  | [44%, 50%] | [27.04, 27.14] |  |
|  | Inverse RC | 74% | 23.06 | (21.60, 24.75) |
|  |  | [64%, 81%] | [22.87, 23.27] |  |
|  | CC | 96% | 17.84 | (16.59, 19.03) |
|  |  | [94%, 97%] | [17.79, 17.88] |  |
|  | MI | 86% | 26.77 | (25.34, 28.21) |
|  |  | [77%, 91%] | [26.59, 26.97] |  |

CC, complete case; CI, confidence interval; MI, multiple imputation; RC, regression calibration;

**Table A5**. In-sample and out-of-sample estimates of R^2^ (A) and slope estimates of the linear regression of observed values on regression calibration (RC) estimates (B) for the simple, GAMLSS based, direct, and inverse RC models by country and (relative) training sample size.

A)

|  |  | Simple RC | | GAMLSS RC | | Direct RC | | Inverse RC | |
| --- | --- | --- | --- | --- | --- | --- | --- | --- | --- |
| **Country** | **Training sample size** | **In-Sample [5%, 95%]** | **Out-Of-Sample  [5%, 95%]** | **In-Sample [5%, 95%]** | **Out-Of-Sample  [5%, 95%]** | **In-Sample [5%, 95%]** | **Out-Of-Sample  [5%, 95%]** | **In-Sample [5%, 95%]** | **Out-Of-Sample  [5%, 95%]** |
| Denmark | 25% | 37% | 35% | 38% | 36% | 40% | 37% | 39% | 36% |
|  |  | [31%, 43%] | [33%, 38%] | [30%, 45%] | [32%, 39%] | [34%, 46%] | [35%, 40%] | [35%, 45%] | [33%, 38%] |
|  | 50% | 36% | 36% | 38% | 37% | 39% | 38% | 37% | 37% |
|  |  | [33%, 40%] | [33%, 39%] | [34%, 42%] | [33%, 41%] | [35%, 43%] | [35%, 42%] | [34%, 41%] | [34%, 42%] |
| Finland | 25% | 24% | 23% | 24% | 24% | 26% | 25% | 25% | 24% |
|  |  | [20%, 28%] | [22%, 25%] | [20%, 29%] | [21%, 26%] | [22%, 31%] | [24%, 27%] | [21%, 29%] | [22%, 26%] |
|  | 50% | 24% | 24% | 25% | 24% | 26% | 25% | 25% | 24% |
|  |  | [22%, 26%] | [21%, 26%] | [22%, 27%] | [22%, 27%] | [24%, 29%] | [23%, 28%] | [22%, 28%] | [21%, 27%] |
| Netherlands | 25% | 33% | 30% | 33% | 29% | 30% | 27% | 32% | 29% |
|  |  | [27%, 38%] | [26%, 32%] | [25%, 41%] | [23%, 33%] | [23%, 37%] | [21%, 32%] | [20%, 38%] | [21%, 33%] |
|  | 50% | 32% | 30% | 33% | 31% | 32% | 30% | 32% | 30% |
|  |  | [29%, 35%] | [27%, 34%] | [29%, 37%] | [27%, 35%] | [29%, 36%] | [25%, 35%] | [25%, 36%] | [23%, 35%] |
| Sweden | 25% | 36% | 36% | 36% | 36% | 37% | 37% | 37% | 37% |
|  |  | [34%, 38%] | [35%, 36%] | [34%, 39%] | [35%, 37%] | [35%, 39%] | [36%, 38%] | [35%, 39%] | [36%, 37%] |
|  | % | 36% | 36% | 36% | 36% | 37% | 37% | 37% | 37% |
|  |  | [34%, 37%] | [34%, 37%] | [35%, 38%] | [35%, 38%] | [36%, 38%] | [36%, 38%] | [36%, 38%] | [35%, 38%] |
| United Kingdom | 25% | 30% | 30% | 31% | 31% | 31% | 31% | 31% | 31% |
|  |  | [28%, 32%] | [30%, 31%] | [30%, 33%] | [31%, 32%] | [29%, 33%] | [31%, 32%] | [29%, 33%] | [30%, 32%] |
|  | 50% | 30% | 30% | 31% | 31% | 31% | 31% | 31% | 31% |
|  |  | [29%, 31%] | [29%, 31%] | [30%, 33%] | [30%, 33%] | [30%, 32%] | [30%, 33%] | [30%, 32%] | [30%, 32%] |

B)

|  |  | Simple RC | | GAMLSS RC | | Direct RC | | Inverse RC | |
| --- | --- | --- | --- | --- | --- | --- | --- | --- | --- |
| **Country** | **Training model size** | **In-Sample [5%, 95%]** | **Out-Of-Sample  [5%, 95%]** | **In-Sample [5%, 95%]** | **Out-Of-Sample  [5%, 95%]** | **In-Sample [5%, 95%]** | **Out-Of-Sample  [5%, 95%]** | **In-Sample [5%, 95%]** | **Out-Of-Sample  [5%, 95%]** |
| Denmark | 25% | 1.00 | 0.99 | 0.83 | 0.80 | 0.99 | 0.96 | 0.92 | 0.90 |
|  |  | [1.00, 1.00] | [0.84, 1.14] | [0.66, 0.97] | [0.60, 0.98] | [0.93, 1.03] | [0.81, 1.11] | [0.81, 1.03] | [0.74, 1.09] |
|  | 50% | 1.00 | 1.00 | 0.83 | 0.83 | 0.99 | 0.98 | 0.96 | 0.96 |
|  |  | [1.00, 1.00] | [0.87, 1.13] | [0.74, 0.93] | [0.69, 0.96] | [0.97, 1.02] | [0.85, 1.12] | [0.90, 1.01] | [0.84, 1.05] |
| Finland | 25% | 1.00 | 1.00 | 0.68 | 0.67 | 1.00 | 1.00 | 1.16 | 1.15 |
|  |  | [1.00, 1.00] | [0.87, 1.13] | [0.52, 0.82] | [0.51, 0.82] | [0.97, 1.05] | [0.89, 1.11] | [1.06, 1.26] | [1.00, 1.29] |
|  | 50% | 1.00 | 1.00 | 0.69 | 0.69 | 1.00 | 1.00 | 1.19 | 1.19 |
|  |  | [1.00, 1.00] | [0.88, 1.12] | [0.61, 0.78] | [0.58, 0.80] | [0.98, 1.03] | [0.90, 1.10] | [1.13, 1.26] | [1.04, 1.31] |
| Netherlands | 25% | 1.00 | 0.97 | 0.88 | 0.83 | 0.64 | 0.61 | 0.98 | 0.90 |
|  |  | [1.00, 1.00] | [0.76, 1.15] | [0.59, 1.02] | [0.54, 1.04] | [0.54, 0.75] | [0.41, 0.82] | [0.63, 1.12] | [0.62, 1.12] |
|  | 50% | 1.00 | 0.98 | 0.88 | 0.86 | 0.84 | 0.80 | 1.00 | 0.96 |
|  |  | [1.00, 1.00] | [0.82, 1.16] | [0.76, 0.98] | [0.67, 1.03] | [0.77, 0.90] | [0.61, 0.99] | [0.77, 1.10] | [0.70, 1.16] |
| Sweden | 25% | 1.00 | 1.00 | 0.71 | 0.70 | 1.00 | 0.99 | 1.14 | 1.13 |
|  |  | [1.00, 1.00] | [0.95, 1.05] | [0.63, 0.78] | [0.62, 0.78] | [0.99, 1.01] | [0.95, 1.04] | [1.11, 1.16] | [1.08, 1.19] |
|  | 50% | 1.00 | 1.00 | 0.71 | 0.71 | 1.00 | 1.00 | 1.13 | 1.13 |
|  |  | [1.00, 1.00] | [0.96, 1.04] | [0.67, 0.75] | [0.66, 0.75] | [0.99, 1.00] | [0.95, 1.04] | [1.12, 1.15] | [1.08, 1.17] |
| United Kingdom | 25% | 1.00 | 1.00 | 0.88 | 0.88 | 0.99 | 0.98 | 1.08 | 1.08 |
|  |  | [1.00, 1.00] | [0.95, 1.04] | [0.85, 0.91] | [0.83, 0.93] | [0.98, 1.00] | [0.94, 1.02] | [1.06, 1.10] | [1.03, 1.12] |
|  | 50% | 1.00 | 1.00 | 0.88 | 0.88 | 0.99 | 0.99 | 1.08 | 1.08 |
|  |  | [1.00, 1.00] | [0.96, 1.04] | [0.86, 0.90] | [0.84, 0.92] | [0.98, 1.00] | [0.95, 1.02] | [1.07, 1.10] | [1.04, 1.12] |

**Table A6**. Country-specific regression calibrated estimates (minutes/week) for the simple RC model based on outgoing call duration, and outgoing and incoming call duration combined, by category of self-report.

|  |  | **Outgoing call duration** | **Outgoing and Incoming call duration combined** |
| --- | --- | --- | --- |
| **Country** | **Self-report categories** | **Estimates (a)** | **Estimates** |
| Denmark | Less than 5 minutes per week | 17.33 | 14.74 |
|  | 5-29 minutes per week | 43.31 | 39.55 |
|  | 30-59 minutes per week | 99.97 | 88.00 |
|  | 1-3 hours per week | 196.44 | 165.99 |
|  | 4-6 hours per week | 302.38 | 250.42 |
|  | More than 6 hours per week | 407.50 | 318.76 |
| Finland | Less than 5 minutes per week | 20.28 | 21.35 |
|  | 5-29 minutes per week | 52.92 | 53.30 |
|  | 30-59 minutes per week | 90.59 | 87.59 |
|  | 1-3 hours per week | 151.00 | 140.57 |
|  | 4-6 hours per week | 228.04 | 199.45 |
|  | More than 6 hours per week | 351.89 | 287.93 |
| Netherlands | Less than 5 minutes per week | 13.55 | 12.43 |
|  | 5-29 minutes per week | 30.24 | 25.45 |
|  | 30-59 minutes per week | 55.43 | 45.62 |
|  | 1-3 hours per week | 81.94 | 66.10 |
|  | 4-6 hours per week | 115.14 | 93.11 |
|  | More than 6 hours per week | 137.58 | 110.99 |
| Sweden | Less than 5 minutes per week | 17.11 | 17.04 |
|  | 5-29 minutes per week | 44.72 | 45.87 |
|  | 30-59 minutes per week | 112.53 | 110.41 |
|  | 1-3 hours per week | 192.00 | 183.64 |
|  | 4-6 hours per week | 288.44 | 266.35 |
|  | More than 6 hours per week | 439.90 | 391.69 |
| United Kingdom | Less than 5 minutes per week | 15.10 | 13.32 |
|  | 5-29 minutes per week | 39.08 | 33.30 |
|  | 30-59 minutes per week | 77.72 | 65.15 |
|  | 1-3 hours per week | 139.05 | 113.47 |
|  | 4-6 hours per week | 222.03 | 179.12 |
|  | More than 6 hours per week | 346.90 | 270.27 |
| United Kingdom: up to 3 phones matched (b) | Less than 5 minutes per week | 15.11 | 13.32 |
|  | 5-29 minutes per week | 39.12 | 33.32 |
|  | 30-59 minutes per week | 77.74 | 65.16 |
|  | 1-3 hours per week | 139.16 | 113.52 |
|  | 4-6 hours per week | 222.35 | 179.44 |
|  | More than 6 hours per week | 347.30 | 270.46 |

(a) the reported estimates for outgoing call duration reflect the ratio [(outgoing call duration/incoming call duration) = 1]

(b) participants indicated two or more numbers and the first three were matched (n= 62018)

**Figure A1.** Parameter estimates for the categorical predictors of the exposure models for the direct (A) and inverse (B) regression calibration (RC) approaches and for the structural measurement error model of the inverse RC approach (C) by country.

Categorical predictors: sex (female versus male), marital Status (living apart and not in a relation versus living together); employment status (inactive versus active), and educational level (elementary school only versus secondary school or higher).

| A  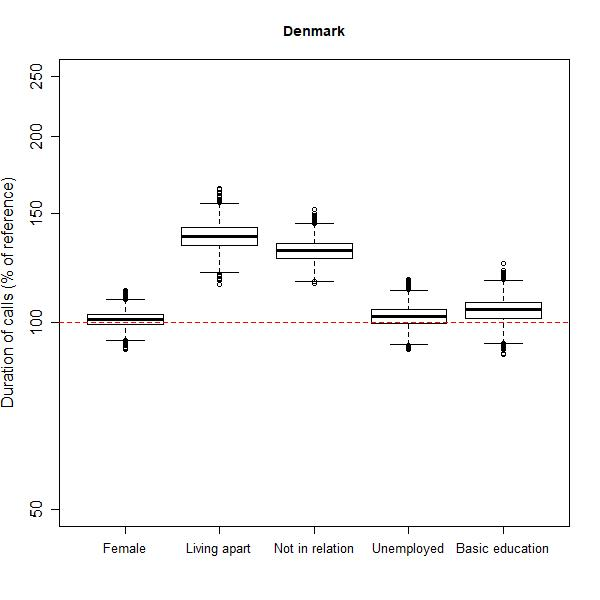 | B  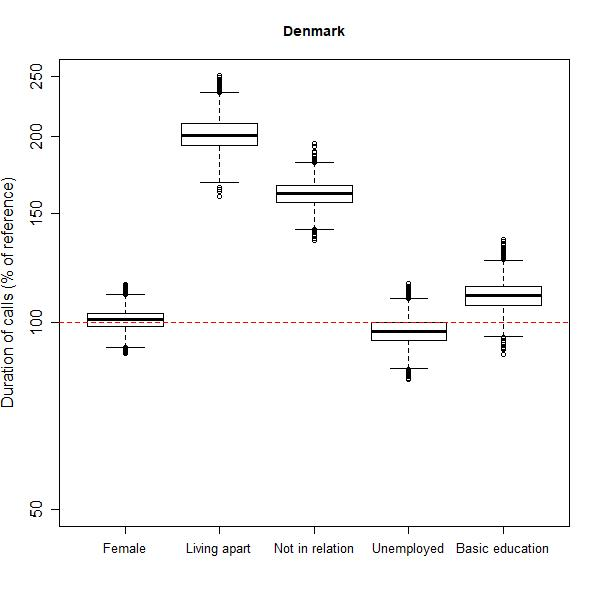 | C  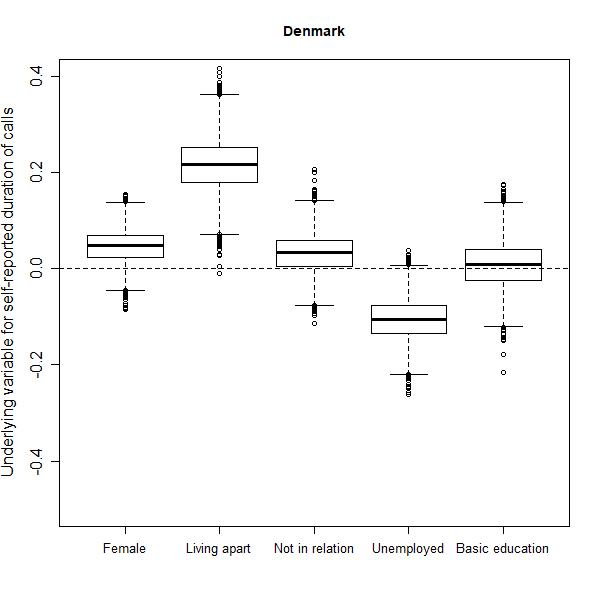 |
| --- | --- | --- |
| 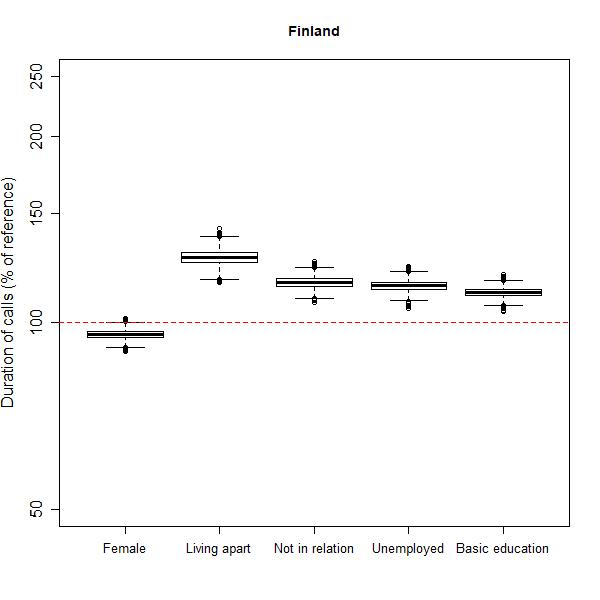 | 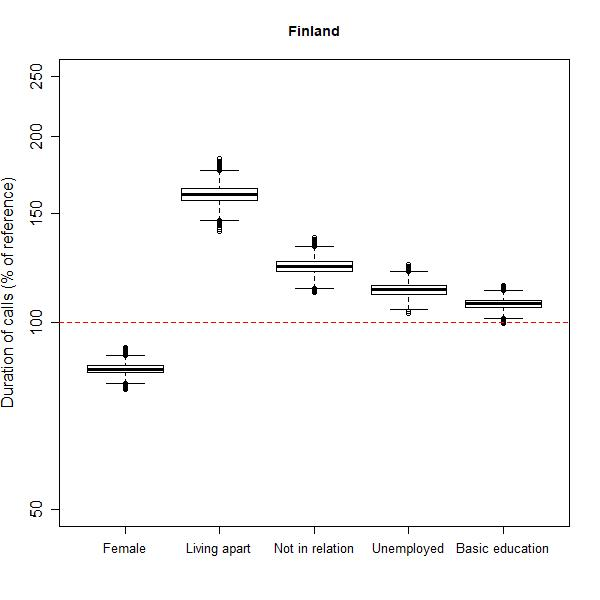 | 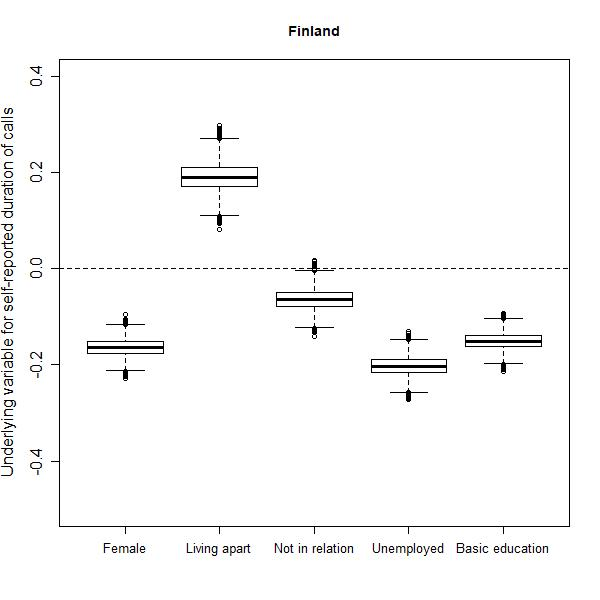 |
| 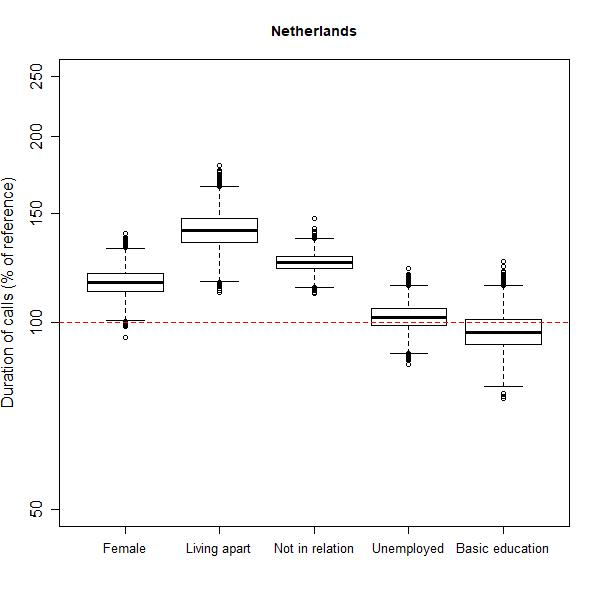 | 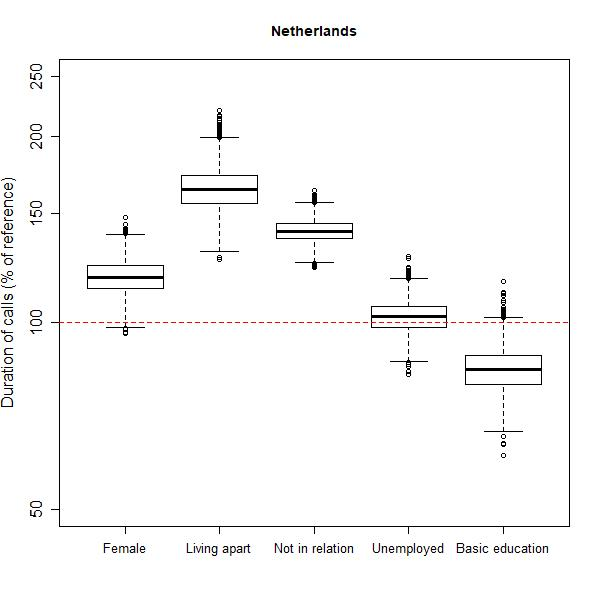 | 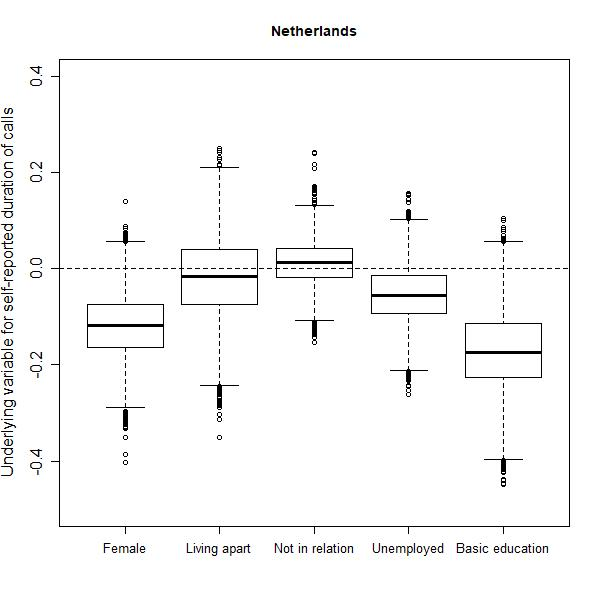 |
| 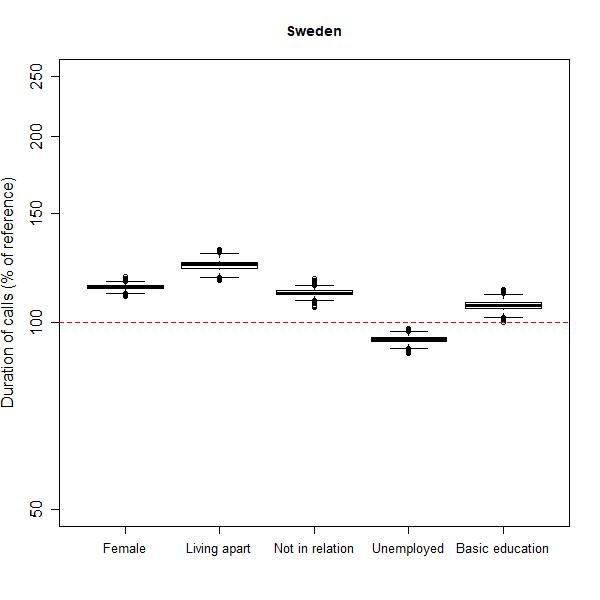 | 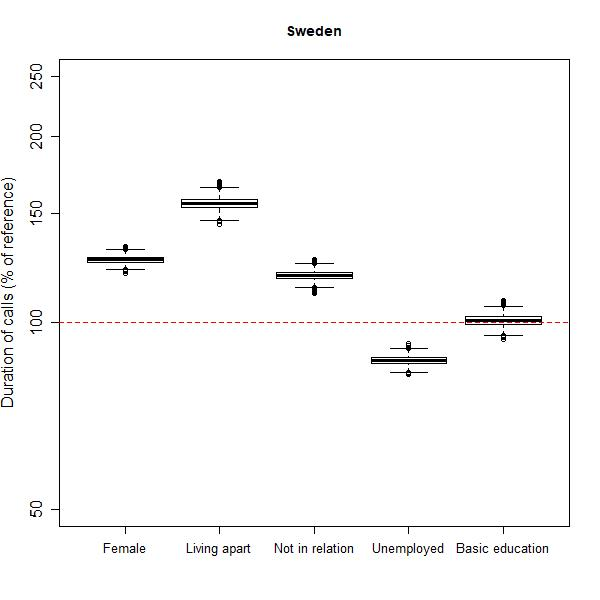 | 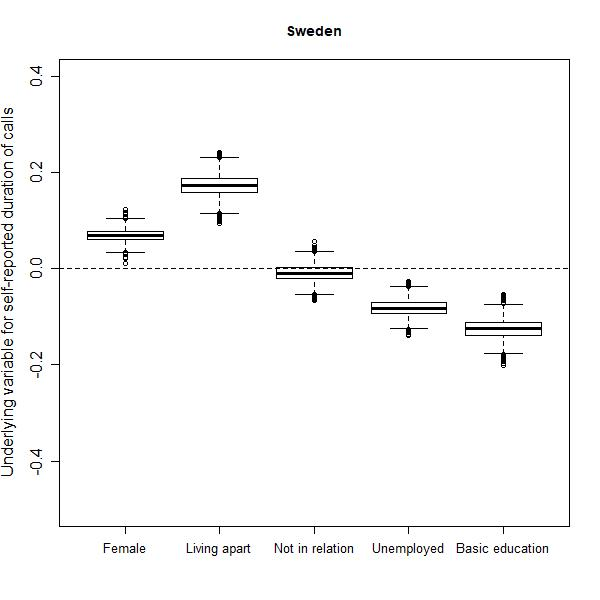 |
| 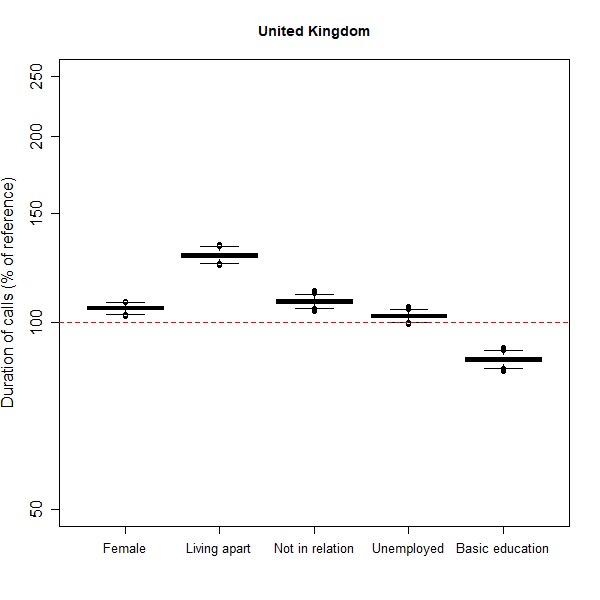 | 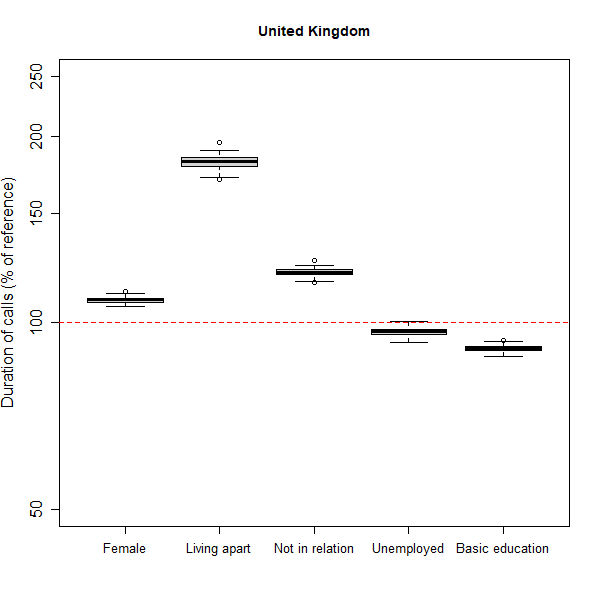 | 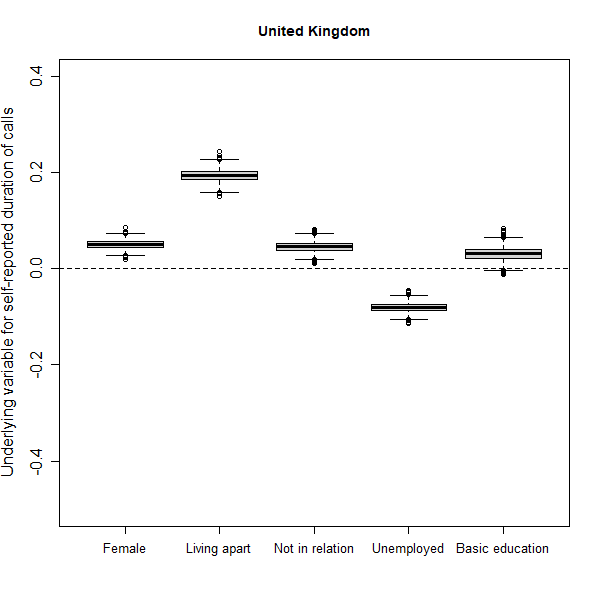 |

**Figure A2**. Effect estimates for age in the exposure models of the direct (A) and inverse (B) regression calibration (RC) approaches, and for age (C) and RECORD (D) in the structural measurement error model of the inverse RC approach by country.

| A  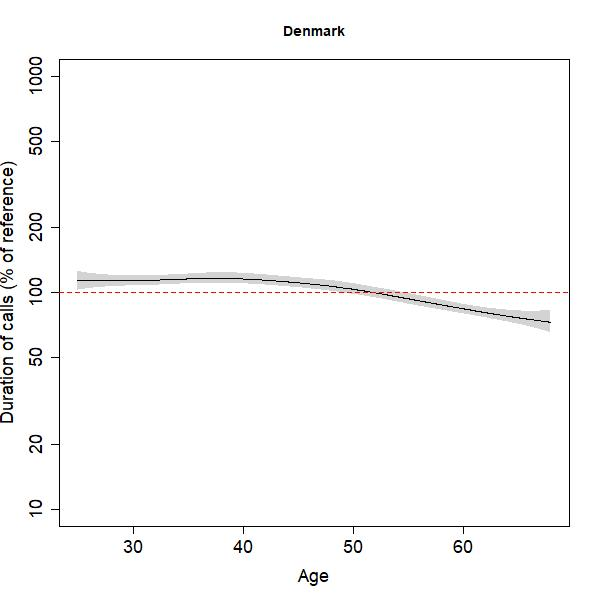 | B  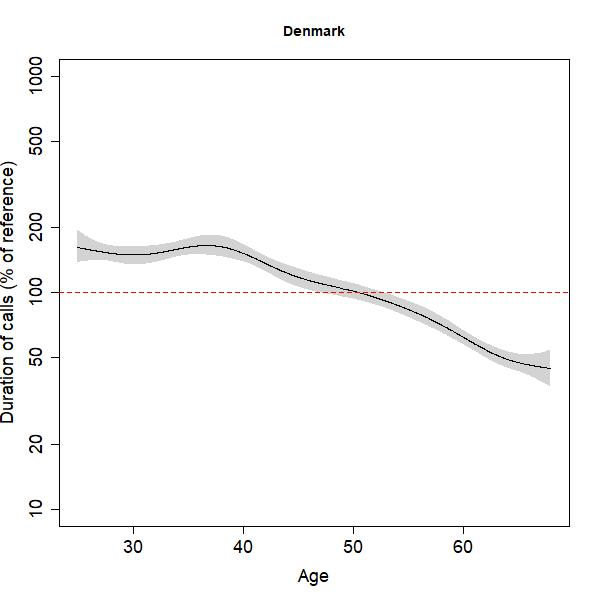 | C  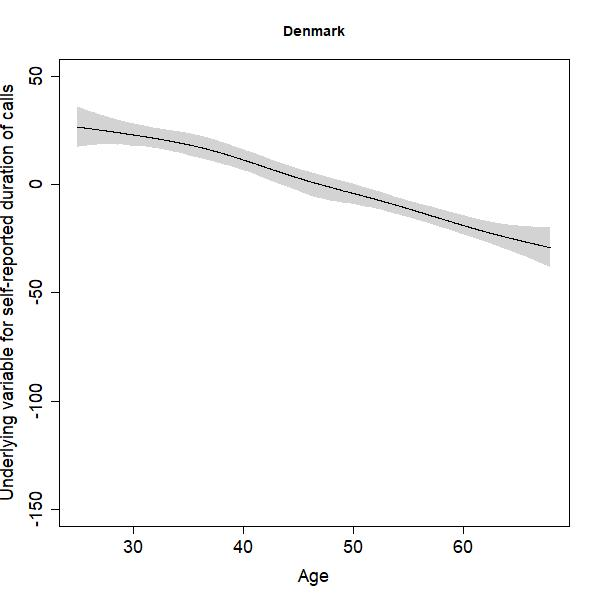 | D  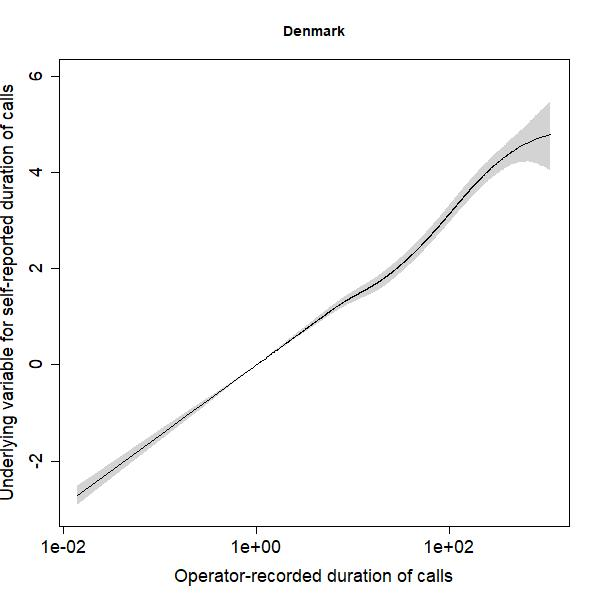 |
| --- | --- | --- | --- |
| 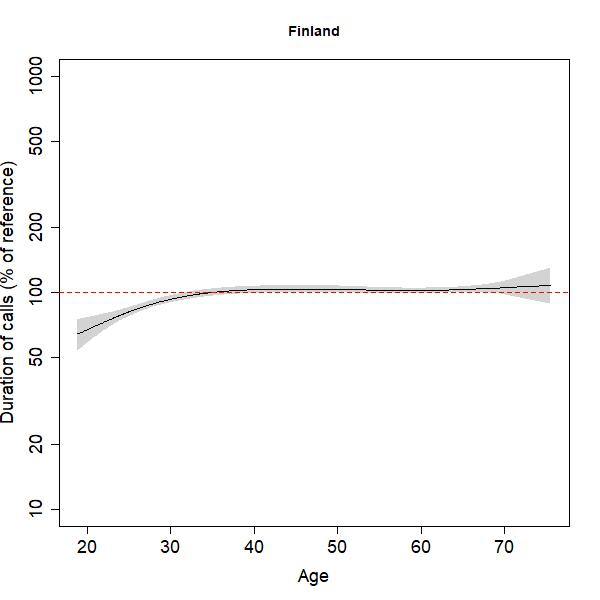 | 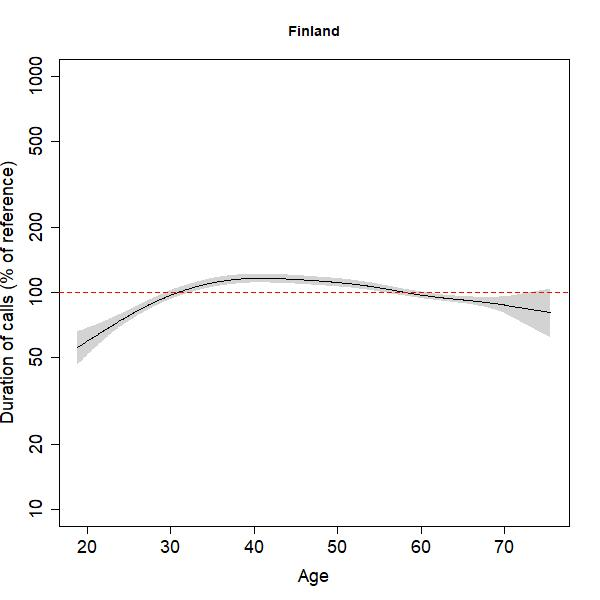 | 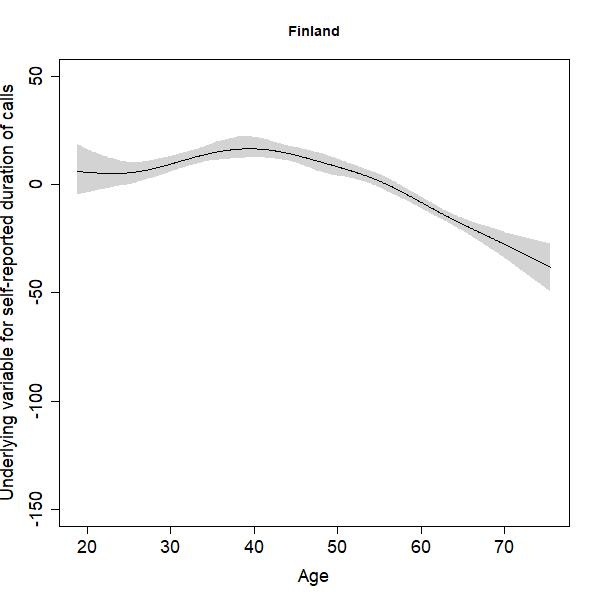 | 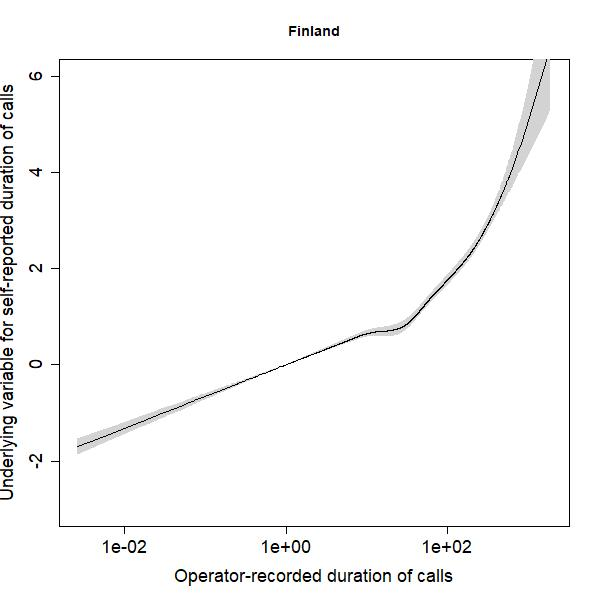 |
| 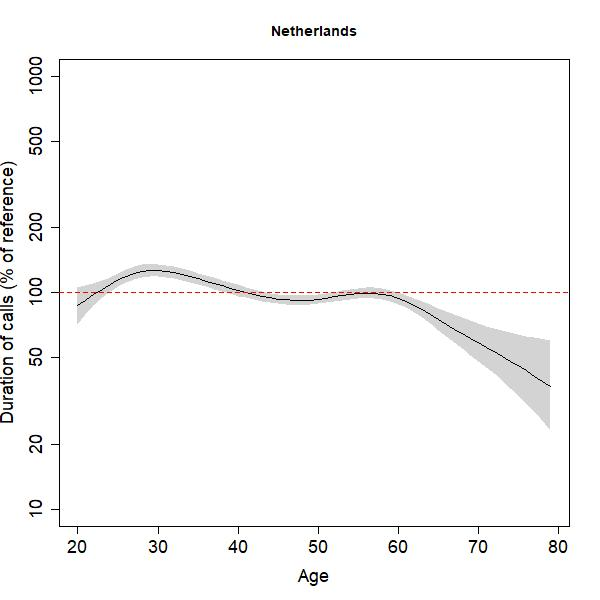 | 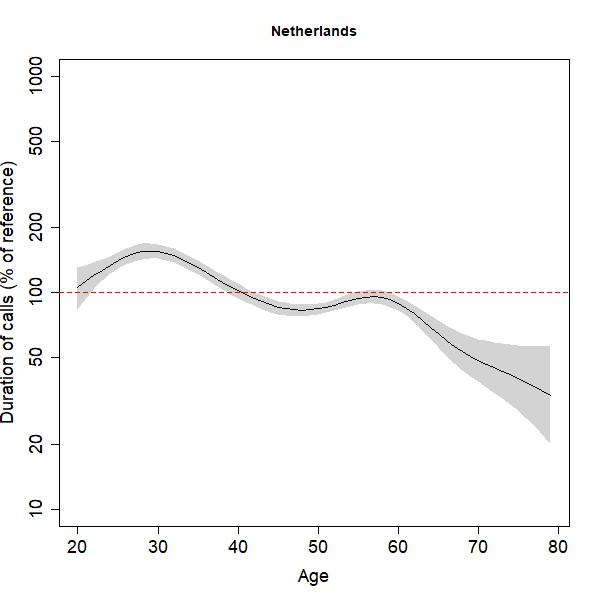 | 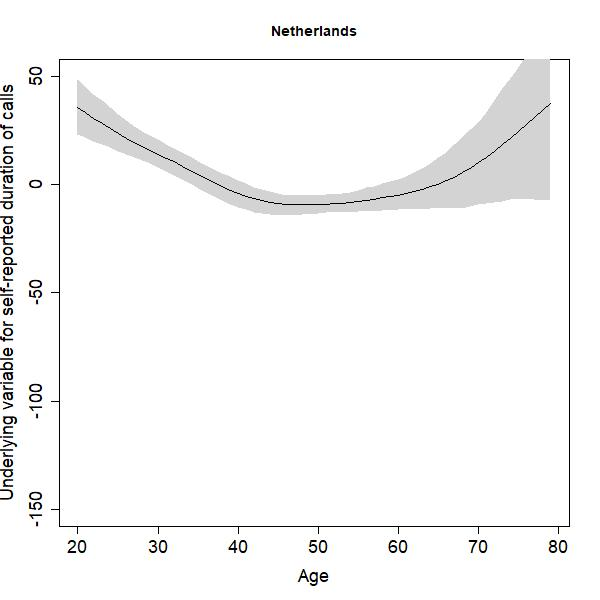 | 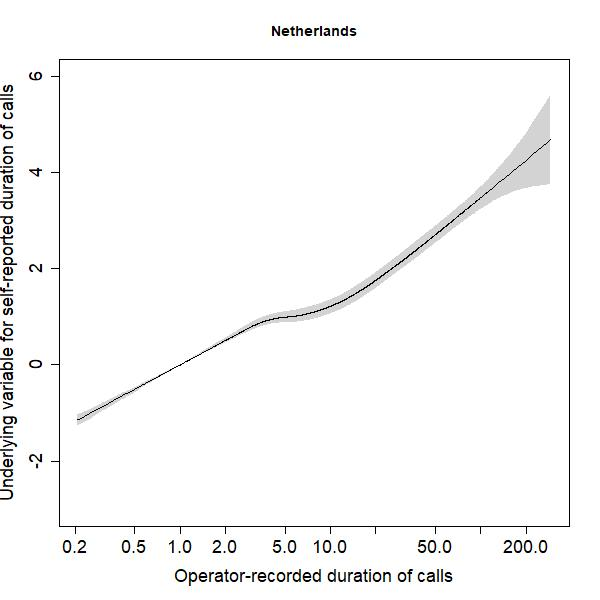 |
| 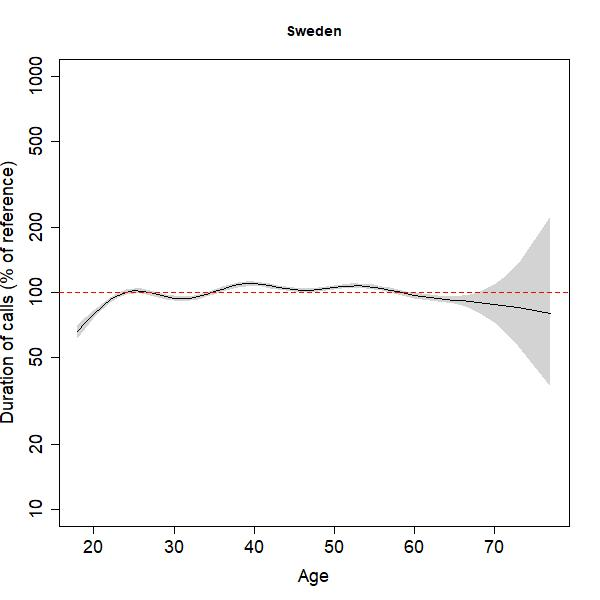 | 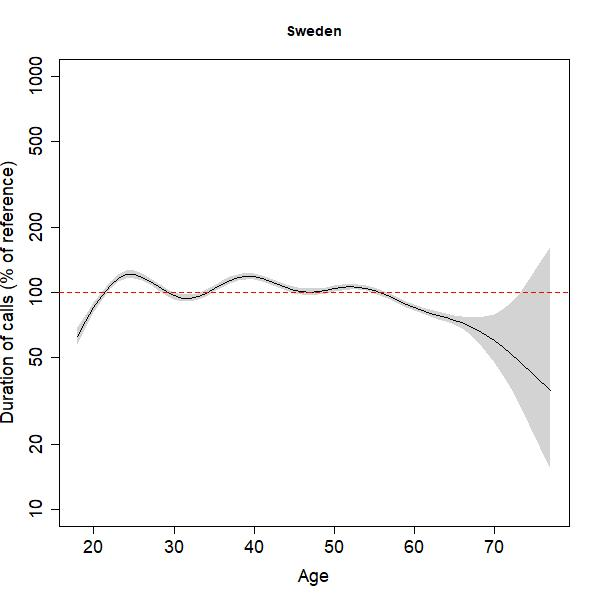 | 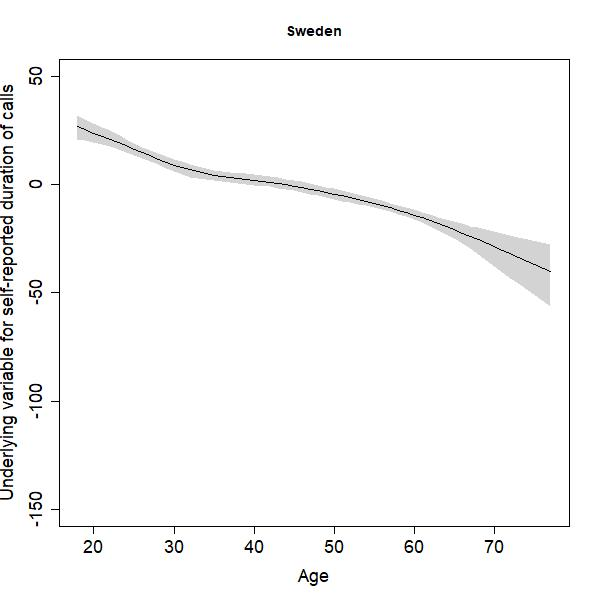 | 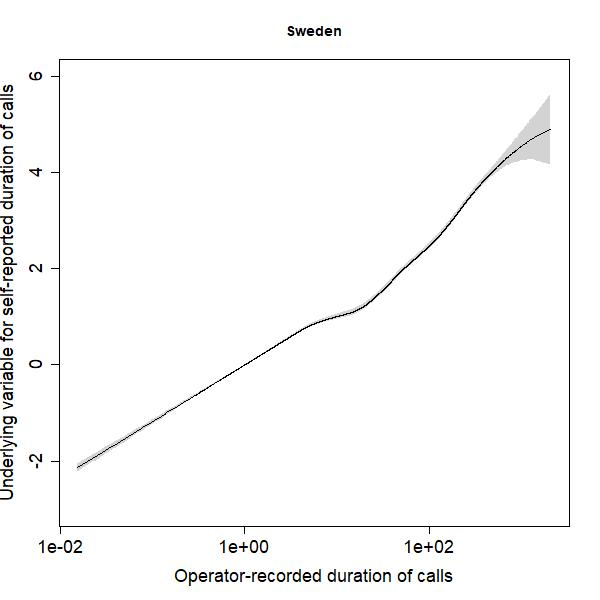 |
| 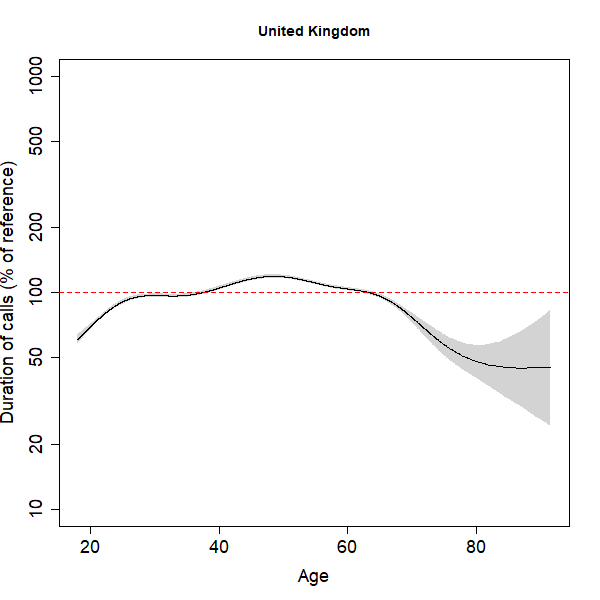 | 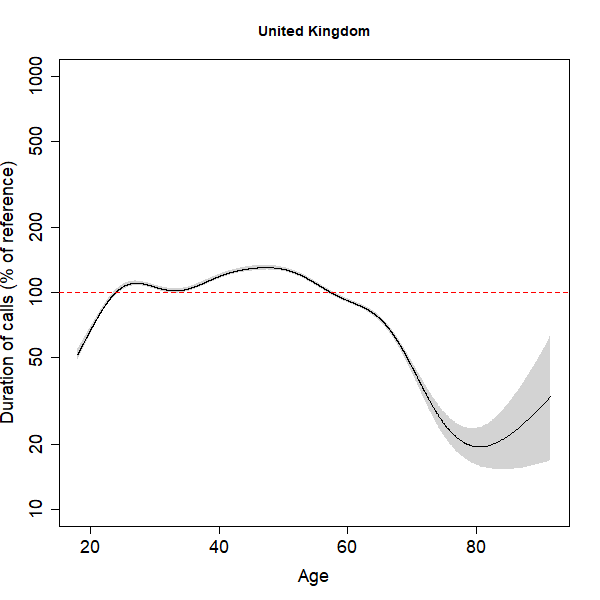 | 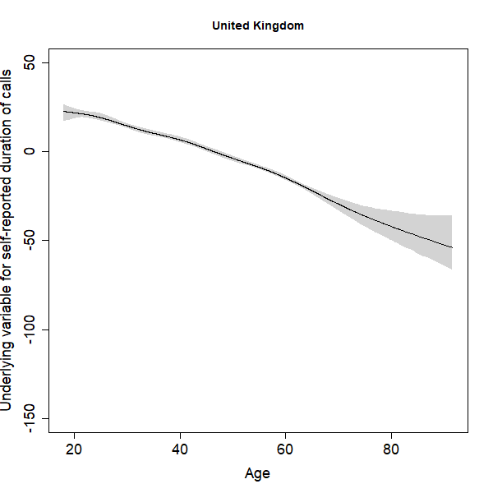 | 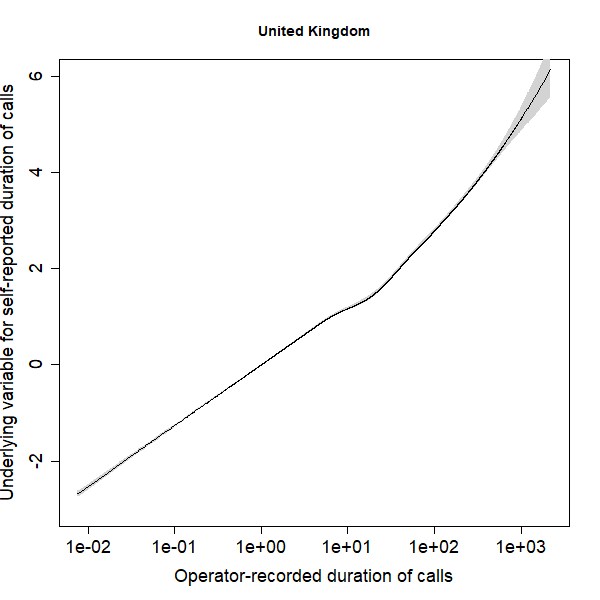 |

**Figure A3.** Observed versus predicted duration of outgoing calls by country for the simple (A) and gamlss-based (B) regression calibration approaches. The regression line was estimated allowing for a non-linear relation between observed and predicted values and allowing the (residual) variance in observed durations to depend on predicted duration using penalized splines (P-splines) as implemented in the gamlss software[8]. Note the different horizontal and vertical scales for the Netherlands.

| A 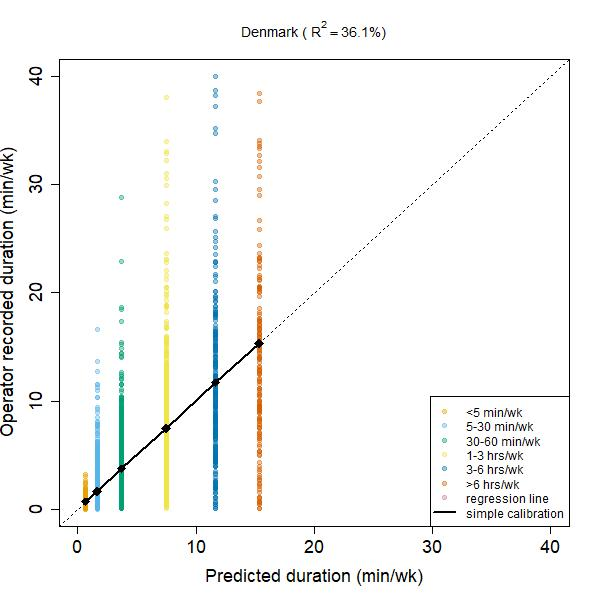 | B  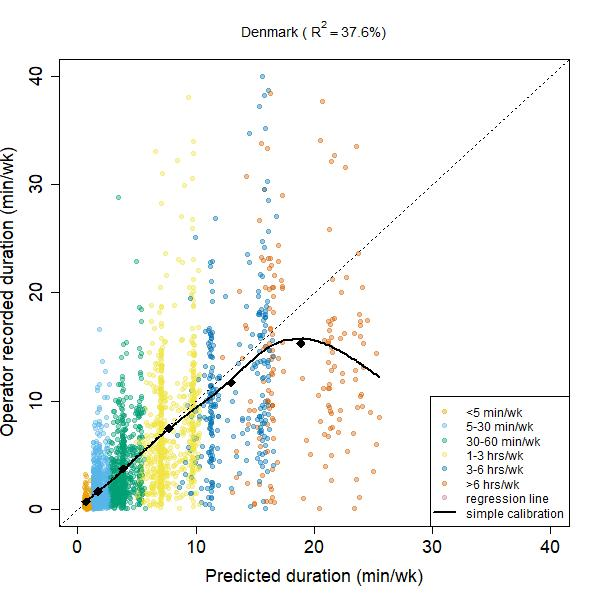 |
| --- | --- |
| 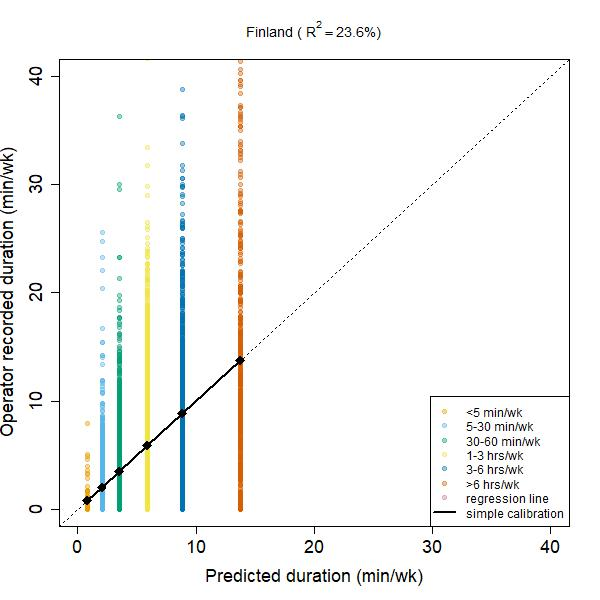 | 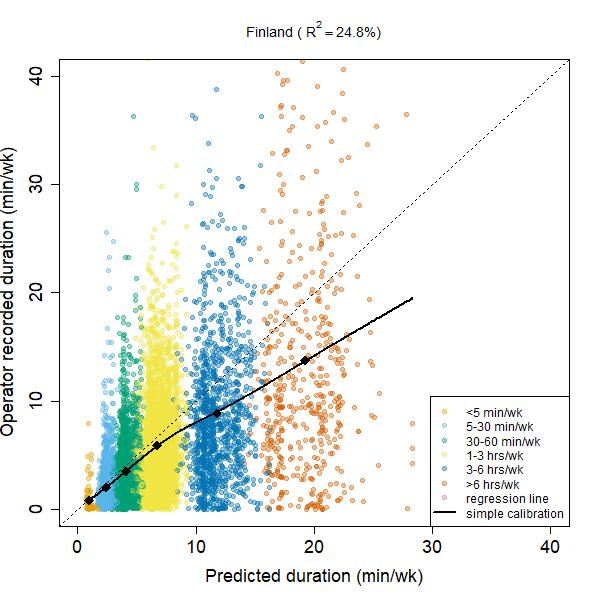 |
| 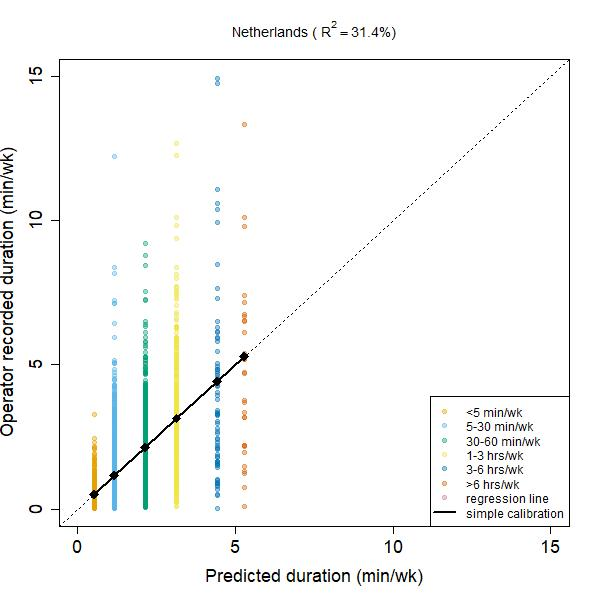 | 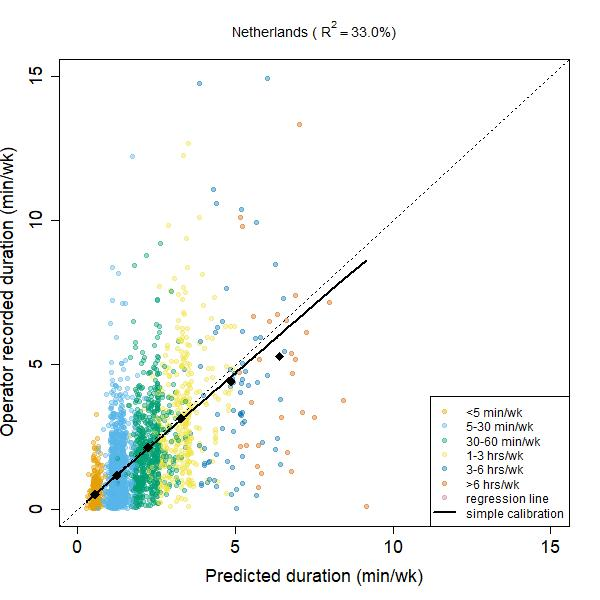 |
| 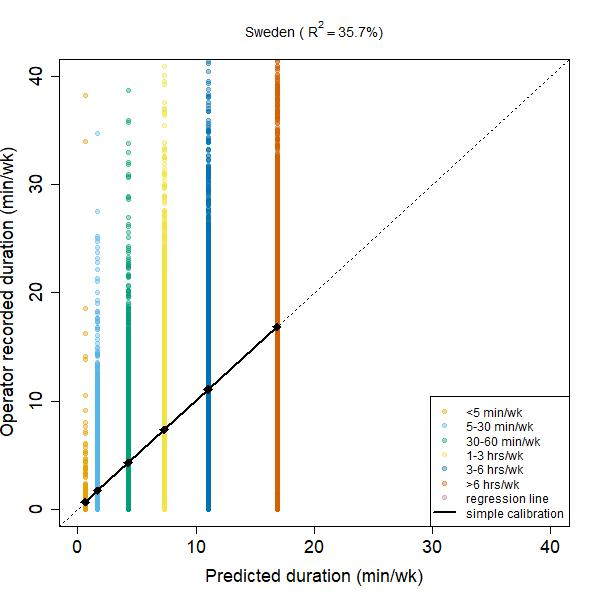 | 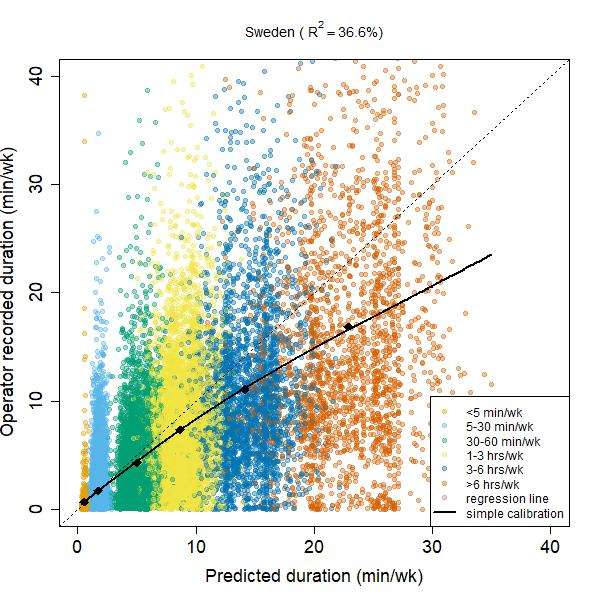 |
| 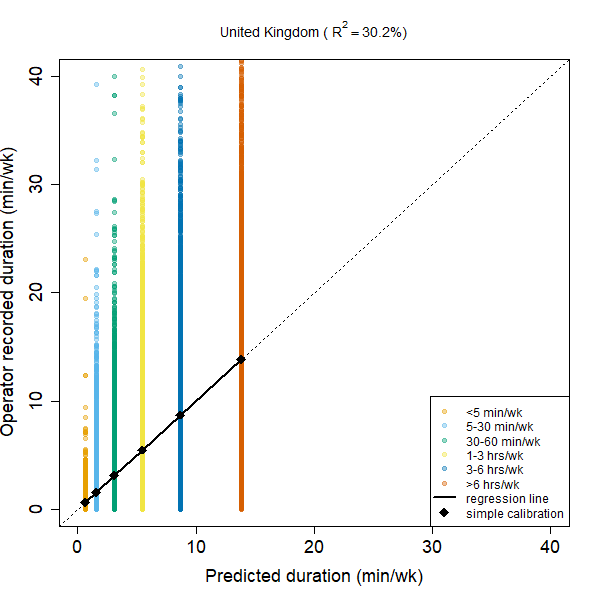 | 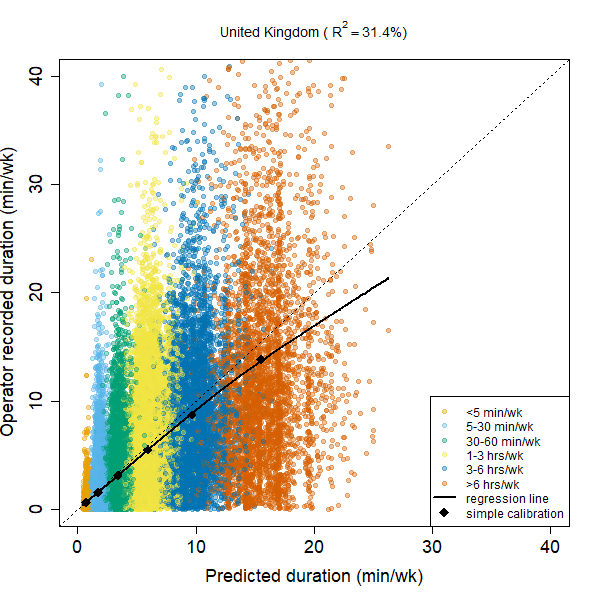 |
